# Direct and indirect mortality impacts of the COVID-19 pandemic in the US, March 2020-April 2021

**DOI:** 10.1101/2022.02.10.22270721

**Authors:** Wha-Eum Lee, Sang Woo Park, Daniel M Weinberger, Donald Olson, Lone Simonsen, Bryan T. Grenfell, Cécile Viboud

## Abstract

Excess mortality studies provide crucial information regarding the health burden of pandemics and other large-scale events. Here, we used time series approaches to separate the direct contribution of SARS-CoV-2 infections on mortality from the indirect consequences of pandemic interventions and behavior changes in the United States. We estimated deaths occurring in excess of seasonal baselines stratified by state, age, week and cause (all causes, COVID-19 and respiratory diseases, Alzheimer’s disease, cancer, cerebrovascular disease, diabetes, heart disease, and external causes, including suicides, opioids, accidents) from March 1, 2020 to April 30, 2021. Our estimates of COVID-19 excess deaths were highly correlated with SARS-CoV-2 serology, lending support to our approach. Over the study period, we estimate an excess of 666,000 (95% Confidence Interval (CI) 556000, 774000) all-cause deaths, of which 90% could be attributed to the direct impact of SARS-CoV-2 infection, and 78% were reflected in official COVID-19 statistics. Mortality from all disease conditions rose during the pandemic, except for cancer. The largest direct impacts of the pandemic were seen in mortality from diabetes, Alzheimer’s, and heart diseases, and in age groups over 65 years. In contrast, the largest indirect consequences of the pandemic were seen in deaths from external causes, which increased by 45,300 (95% CI 30,800, 59,500) and were statistically linked to the intensity of non-pharmaceutical interventions. Within this category, increases were most pronounced in mortality from accidents and injuries, drug overdoses, and assaults and homicides, while the rate of death from suicides remained stable. Younger age groups suffered the brunt of these indirect effects. Overall, on a national scale, the largest consequences of the COVID-19 pandemic are attributable to the direct impact of SARS-CoV-2 infections; yet, the secondary impacts dominate among younger age groups, in periods of stricter interventions, and in mortality from external causes. Further research on the drivers of indirect mortality is warranted to optimize interventions in future pandemics.

## Introduction

As the official death toll of the coronavirus disease 2019 (COVID-19) continues to grow, the full impacts of the pandemic on a range of conditions remain debated. In the United States (US), over 74 million confirmed cases and 887,000 official deaths were reported as of February 1, 2022 (Johns Hopkins University, 2022). A pandemic of the magnitude of COVID-19 has secondary effects on unrelated health conditions; for instance, non-COVID-19 deaths increased in Spring 2020 at the height of the first wave in part due to avoidance of the healthcare system (Sharma et al., 2021; Woolf et al., 2020).

Excess mortality approaches have been used for over a century to capture the full scope of large-scale infectious disease events, heatwaves, and earthquakes, by measuring the rise in mortality over a historical baseline (Serfling, 1963; Weinberger et al., 2020). In the early phase of the pandemic, these approaches highlighted substantial underestimation in official statistics of COVID-19 deaths due to limited viral testing (Weinberger et al., 2020). More recent analyses have examined excess mortality patterns for specific causes of death by age and socio-demographic groups and have compared the COVID-19 death toll between countries (Banerjee et al., 2020, p. 19; Islam et al., 2021; Karlinsky & Kobak, 2021; Mena et al., 2021; Rossen, 2021; Woolf et al., 2021). Yet, it remains a challenge to separate the direct and indirect impacts of the pandemic. The direct impacts of the virus include potential effects on deaths ascribed to chronic conditions; for instance, death in a diabetic patient could have been triggered by SARS-CoV-2 infection and be coded as diabetes. The indirect impacts of the pandemic include avoidance of the healthcare system for treatment of acute conditions and for management of underlying chronic conditions, stressed healthcare systems in period of high COVID19 incidence, and societal disruptions (Sharma et al., 2021). In particular, the indirect impacts of the pandemic on mental health, violence, and addiction remain debated, with potentially large impacts on mortality (Faust, Du, et al., 2021; Faust, Shah, et al., 2021).

There is substantial heterogeneity in the trajectory of the COVID-19 pandemic and public health measures across the US, providing an opportunity to separate the contributions of viral infection on mortality from that of pandemic interventions, in a large country with homogenous death ascertainment. Here, we use time series approaches to separate the direct consequences of SARS-CoV-2 infection on age-state- and cause-specific mortality from the indirect effects of the pandemic. Our analyses cover three large waves of infections from March 2020 to April, 2021 in the US. We also compare our excess mortality estimates with serology (Asten et al., 2021; Verity et al., 2020) and explore between-state variation in SARS-CoV-2 infection fatality rates. Our analyses contrast the direct and indirect effects of the pandemic on several chronic conditions and shed light on the long-lasting consequences on violence and overdoses.

## Data and Methods

### Mortality Data

We obtained weekly mortality counts from the National Center for Health Statistics (NCHS) for the period August 1, 2014– April 30, 2021; we included 2014-2019 data to construct robust historical model baselines (Centers for Disease Control and Prevention, 2021a, 2021b). Data were stratified by state, 6 age groups (all ages, under 25 years, 25–44, 45–64, 65–74, 75–84, and over 85), and 8 underlying mortality causes (all causes, respiratory conditions, Alzheimer’s disease, cancer, cerebrovascular diseases, diabetes, heart disease, external causes; see supplement for disease codes). External causes include suicides, accidents, homicides, and opioids, among other conditions. We used aggregated mortality counts ascribed to COVID-19, influenza, pneumonia, and chronic lower respiratory diseases as an indicator of ‘respiratory mortality’, which was our most specific indicator of excess deaths directly caused by SARS-CoV-2 infection. We compiled weekly deaths with any mention of COVID-19 anywhere in the death certificate and considered those to be the official COVID-19 statistics (Centers for Disease Control and Prevention, 2021c).

To further explore patterns in external mortality causes, which include a range of conditions, we obtained additional monthly data by subcategories of deaths, including suicides, assaults and homicides, drug overdoses, accidents and unintentional injuries, and motor vehicle accidents (a subset of accidents) (Centers for Disease Control and Prevention, 2021d, 2021e). We also downloaded monthly deaths from external causes combined by age and region (Centers for Disease Control and Prevention, 2021f). External causes of deaths are typically released several months after other conditions and detailed data were unavailable at a weekly resolution.

### Other datasets

Age- and state-specific population estimates were obtained from CDC (Centers for Disease Control and Prevention, 2021g) and used to calculate mortality rates. To validate our excess mortality approach, we compared our mortality estimates against CDC state-specific seroprevalence surveys (Centers for Disease Control and Prevention, 2021h). We used data on the proportion of the population with SARS-CoV-2 antibodies to the nucleocapsid by late April 2021 to compare with our excess death estimates at the end of April 2021, given a similar delay between infection and death and infection and rise in antibodies. As the nucleocapsid is not a component of the vaccines used in the US, the serologic assay captures natural infections. We used our comparison of excess mortality against serology to estimate the infection fatality rate. We ran sensitivity analyses on the maximum seroprevalence reported during the study period, rather than seroprevalence at the end of the study period, to account for potential waning of natural immunity.

To adjust excess mortality models for the contribution of influenza, we downloaded weekly surveillance data from CDC FluView (Rudis et al., 2021). Finally, to evaluate the putative impacts of public health interventions on cause-specific mortality, we compiled the health containment index from the COVID-19 government response tracker which measures the strength of interventions by week and state (Oxford University, 2021).

All data used in the analysis were publicly available and exempt from human subjects review; data and code have been posted in a GitHub repository that will be publicly released upon publication.

### Analytic Approach

#### Excess Mortality Models

Before running mortality models, we adjusted mortality counts for reporting delays using a modified NobBS package in R (McGough et al., 2020). Even though there was little reporting delay for the data presented in this study, we wanted to ensure comparability with prior work (Weinberger et al., 2020). After adjustment, we applied seasonal regression models to weekly mortality rates in the pre-pandemic period, August 1, 2014–March 1, 2020, and estimated the baselines for each mortality cause, age group, and state (Goldstein et al., 2012; Weinberger et al., 2020; and supplement for more details). The model included harmonic terms for seasonality, time trends, and a proxy for weekly influenza incidence. We fitted the model to data until March 1, 2020 and projected the baseline forward until April 30, 2021. We estimated weekly excess mortality by subtracting the predicted baseline from the observed mortality that week; total excess mortality was the sum of weekly excesses (positive or negative) during March 1, 2020 – April 30, 2021. We used block bootstraps to generate 95% prediction intervals and obtain uncertainty intervals on estimates. For context, we also estimated excess mortality for the severe 2017/2018 influenza season.

We ran cause-specific excess mortality analyses nationally and for states that had sufficient mortality counts, as weekly death counts below 10 were blanked due to privacy concerns. States missing more than 2 weeks of data between March 2020 and April 2021 were excluded from the corresponding analyses. We ran respiratory excess mortality analyses for 16 states and non-respiratory analyses for 33 states (see Supplement for full list).

#### Estimation of direct and indirect pandemic impacts

To assess the direct and indirect impacts of the pandemic on mortality, we performed several correlation and regression analyses evaluating the trajectory of different causes of deaths by age and geography, building on earlier work on the 1968 influenza pandemic (Reichert et al., 2004) and COVID-19 (Sharma et al., 2021) (Details in the Supplement). First, we tested whether weekly non-respiratory excess mortality became more correlated with respiratory excess mortality during March 2020-April 2021, compared to the pre-pandemic period. This would signal a direct but undetected effect of COVID-19 on non-respiratory mortality. Second, we assessed whether states that experienced high COVID-19 mortality concomitantly experienced high mortality from other causes during the pandemic. We used our respiratory excess mortality indicator, and the official COVID-19 death toll, as complementary measures of COVID-19 mortality. Third, to quantify the relative contributions of SARS-CoV-2 infection (direct impact) and non-pharmaceutical interventions (indirect impact) on mortality, we regressed weekly cause- and age-specific excess mortality against COVID-19 deaths and the COVID-19 containment index, after exploring different lags between predictors and outcomes. Regression models were run nationally and at the state level. Uncertainty in weekly excess mortality estimates was propagated into the regression models (Supplement).

#### Validation of excess deaths based on serology; estimation of Infection Fatality Rate (IFR)

Since deaths are ultimately the result of infections, serology can provide a validation of excess mortality estimates for mortality indicators that are specific of SARS-CoV-2. To test the validity of the excess mortality approach, we regressed cumulative excess respiratory mortality rates against SARS-CoV-2 seroprevalence estimates at the state level, in a model without intercept since there should be a direct correspondence between rates of infection and death. We repeated this analysis with all-cause excess mortality, official COVID-19 deaths, and excess mortality in individuals over 65 years as the outcome variable. These analyses provided both a statistical validation of our excess mortality approach and an opportunity to estimate the IFR, which is the slope of these regressions. We propagated the errors obtained in excess mortality and seroprevalence estimates into IFR estimates (Supplement).

## Results

### Overall mortality patterns

Across the US from March 1, 2020— April 30, 2021, there were 519,320 deaths officially attributed to COVID-19. During the same period, we estimate 507000 (95% Confidence Intervals (CI) 487000, 526000) excess respiratory deaths and 666,000 (95% CI 556000, 774000) excess deaths due to all-cause (Table 1). National mortality patterns comprise three waves from March 1–June 20, 202; June 21–September 19, 2020; and September 20, 2020 to April 30, 2021, with varying timing and intensity by state (Figure 1A and S1-S8). The first wave was concentrated in Northeastern states, while the Southern and Western states showed larger mortality increases during the second and third waves.

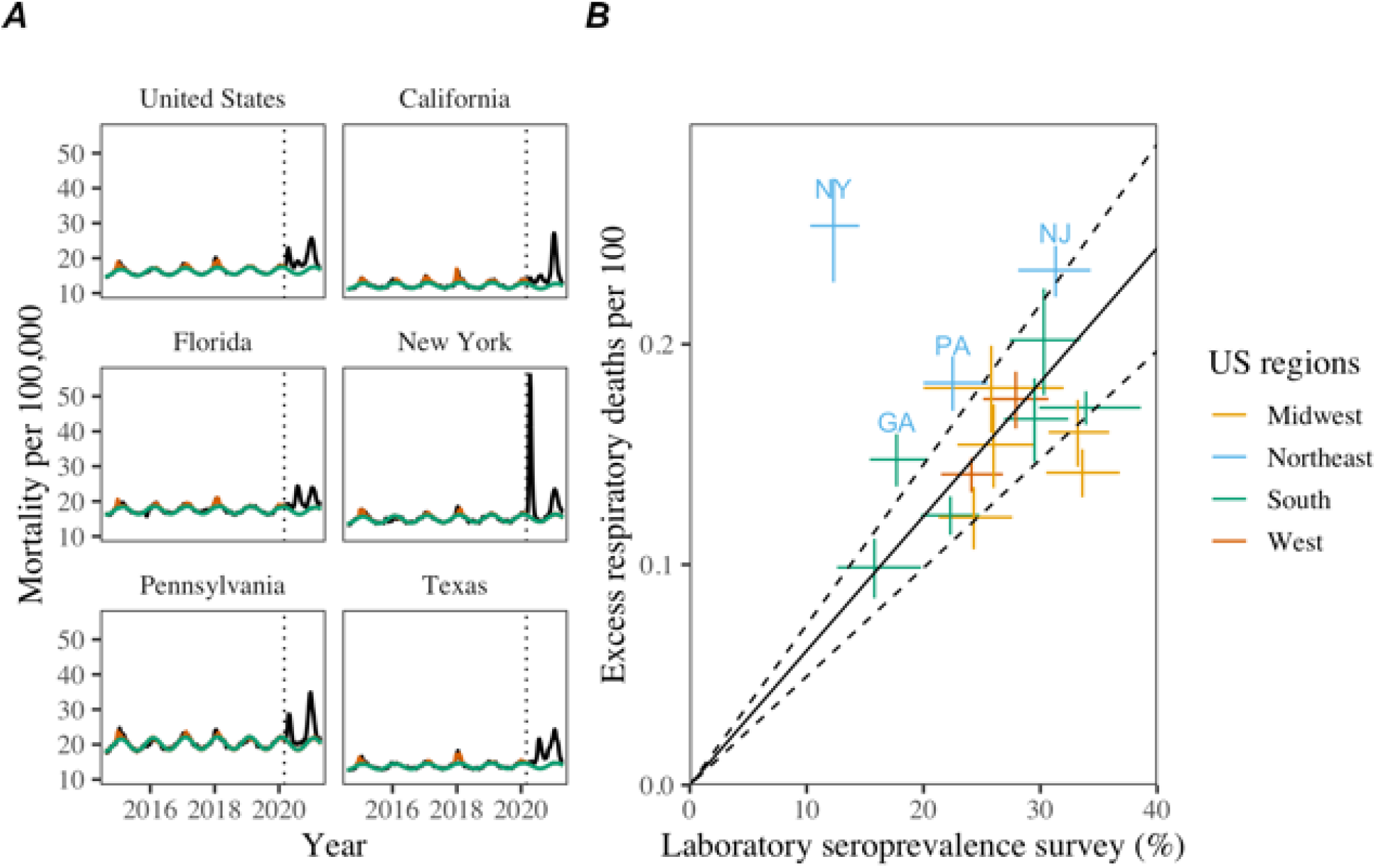

**Table 1.**
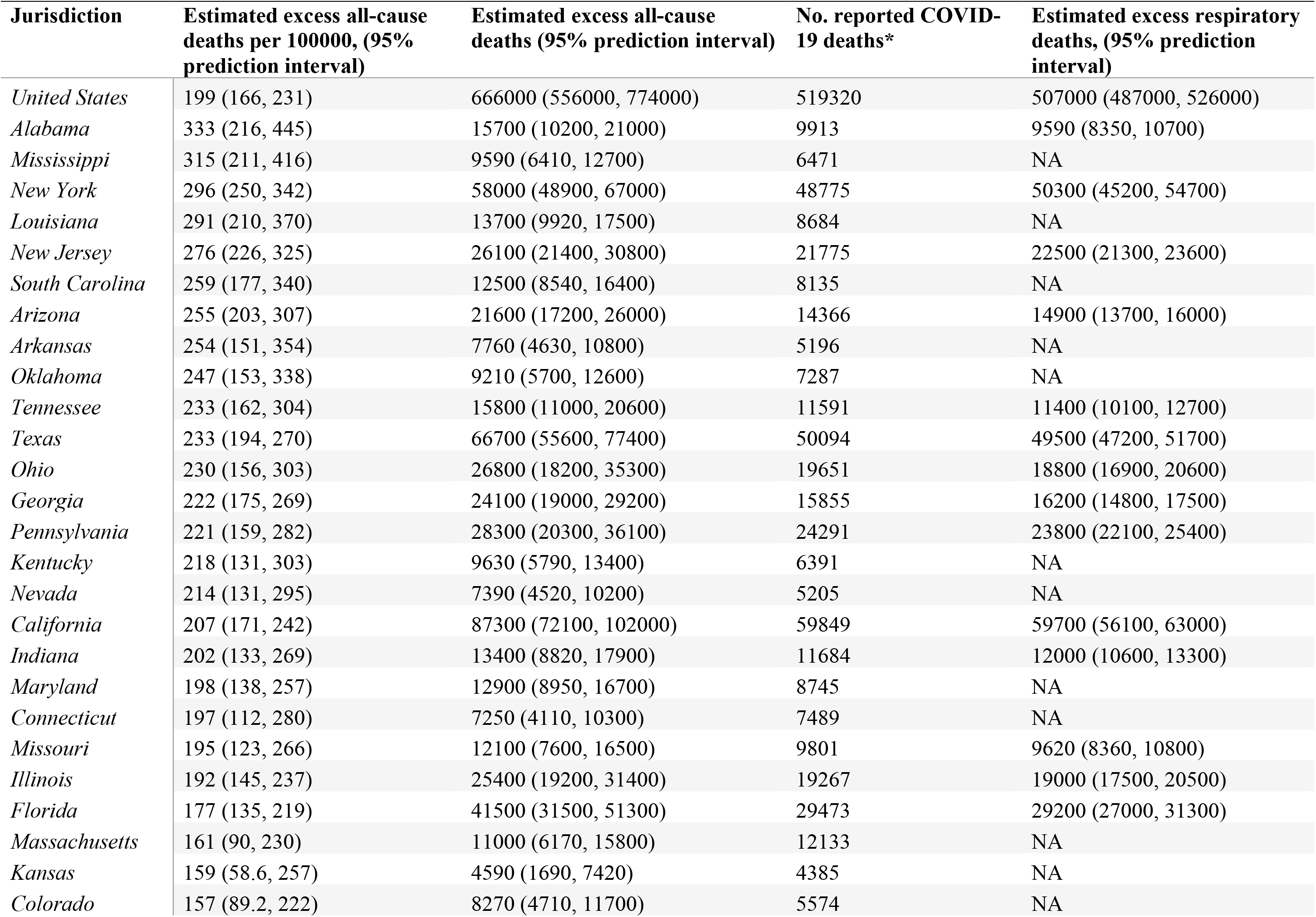
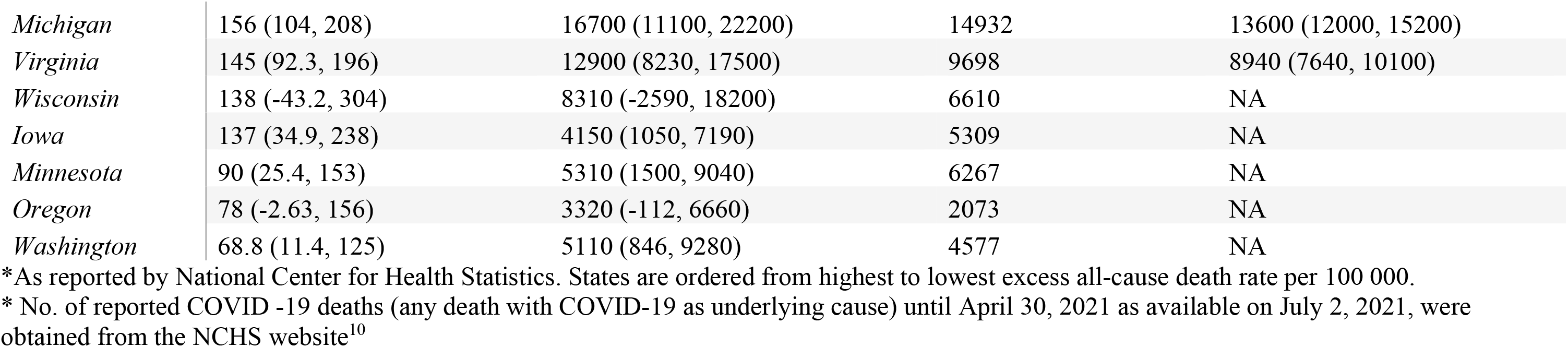
Reported COVID-19 deaths, Compared with Excess Deaths from All-Causes and Respiratory Diseases: March 1, 2020 – April 30, 2021

Excess respiratory mortality showed significant, positive correlation with seroprevalence surveys in each state (Figure 1B). Seroprevalence estimates ranged between 4.7 and 28% at the end of April 2021, with a population-weighted national seroprevalence of 22.8%. New York experienced higher than predicted excess mortality with respect to the reported serologic infection rates. This remained true in sensitivity analyses based on the maximum reported seroprevalence at any time point of the study period. The nationwide infection fatality rate (IFR) was estimated at 0.61% (0.49 - 0.73) based on excess respiratory mortality and 0.86% (0.72 - 0.99) based on all-cause excess mortality (Figure S9). Use of official COVID-19 deaths determined an IFR of 0.72% (0.62 - 0.81); interestingly, official COVID-19 deaths did not align with serology data as well as excess respiratory deaths (Figure S9). The IFR was significantly higher in individuals over 65 years, estimated at 5.7% (4.7-6.8).

By April 2021, the COVID-19 pandemic had caused significantly higher mortality compared to previous seasonal influenza outbreaks in the US. Respiratory excess mortality during COVID-19 exceeded the impact of the severe 2017/2018 influenza season 8.2 times (range across states 4.7-28.3).

### Direct and indirect pandemic impacts by cause of death

Excess mortality increased during the pandemic for 6 of the 7 non-respiratory conditions studied, although the timing and intensity of excess mortality varied by disease (Table 2, Figure 2). Cancer mortality was the only mortality condition that did not increase during the pandemic. Cancer deaths have remained below historic levels since March 2020, although cumulative weekly departures from baseline were not significantly different from zero (Table 2). In contrast, mortality from Alzheimer’s, diabetes, and heart disease rose during the pandemic, with the trajectory of excess mortality matching that of respiratory mortality in the 3 pandemic waves (Figures 2 for national patterns, and Figures S3-S8 for state-specific data). Across these causes of death, the first excess mortality peaks occurred within one week of the first respiratory mortality peak on April 18, 2020, and synchronicity between mortality causes was most evident in New York and New Jersey in the first wave. The third wave had a consistently larger peaks across diseases (Figure S10).

**Figure 2.**
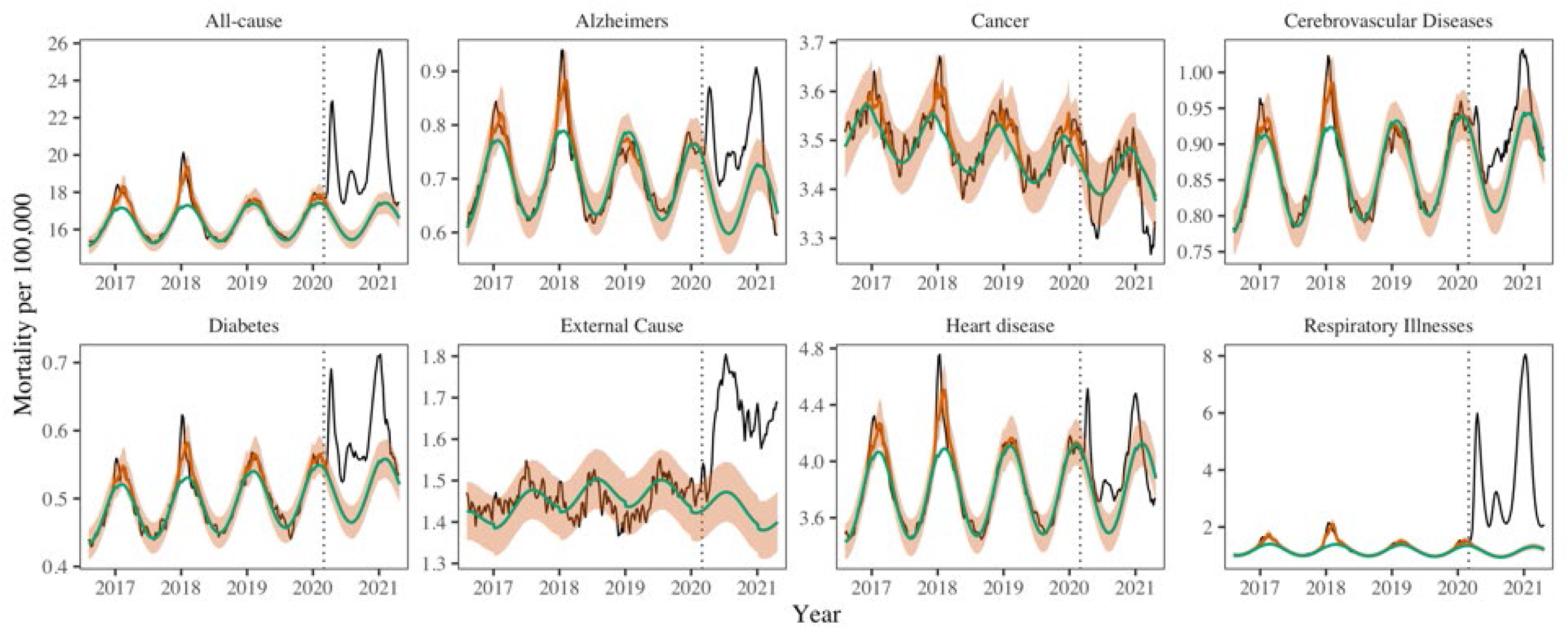
National mortality rate (per 100,000) for 8 causes of death. The black line shows observed data, the green line shows seasonal baseline, the orange shading the 95% CI on the seasonal baseline, and the red line shows model predictions with seasonal variation and influenza circulation. The dotted red lines show the upper and lower 95% confidence intervals. Excess mortality attributed to the COVID-19 pandemic is defined as the area between the black and green line from March 1, 2020. The dotted vertical line marks the start of the pandemic on March 1, 2020.

**Table 2.** Estimation of the direct impacts of COVID-19 on non-respiratory conditions

| **Cause of Death** | **Estimated excess deaths (95% prediction interval)** | **% of excess deaths directly attributable to COVID-19\*** |
| --- | --- | --- |
| All-Cause | 666000 (556000, 774000) | 90 (82, 99) |
| Alzheimer's | 17600 (8800, 26300) | 58 (30, 86) |
| Diabetes | 15800 (10500, 21000) | 59 (42, 76) |
| Cancer | -4800 (-18400, 8600) |  |
| Cerebrovascular Disease | 8500 (1600, 15400) | 50 (18, 83) |
| External Cause | 45300 (30800, 59500) |  |
| Heart Disease | 27800 (-3100, 58000) | 97 (40, 158) |

\* Shown only when COVID-19 is a significant covariate ( $p < 0.05$ ); estimates are derived from a model where weekly excess mortality is regressed against weekly COVID-19 intensity and weekly interventions on a national scale, for the 60 weeks of March 2020 - April 2021.

We found a significant rise in deaths from external causes during the pandemic period March 2020-April 2021, corresponding to 45,300 (30,800, 59,500) excess deaths nationally (Figure 2). The largest excess mortality rates from external causes were estimated in states that also had high baseline death rates from these conditions (Figure S11). The weekly trajectory of mortality from external causes did not align with that of respiratory mortality. We further analyzed subcategories of external causes that were available on a monthly resolution (Figure 3). The largest excess death tolls observed during this period were from accidents and injuries (23,800 (8,400-39,200), an 11% increase over baseline), drug overdoses (15,300 (7,500-23,100), a 15% increase) and assaults and homicides (5,100 (2,700-7,600), a 21% increase, Table 3). Overdoses were the first to peak in May 2020, followed by accidents and assaults in July 2020. In contrast, mortality from suicides remained within historic baselines throughout the COVID-19 period.

**Figure 3:**
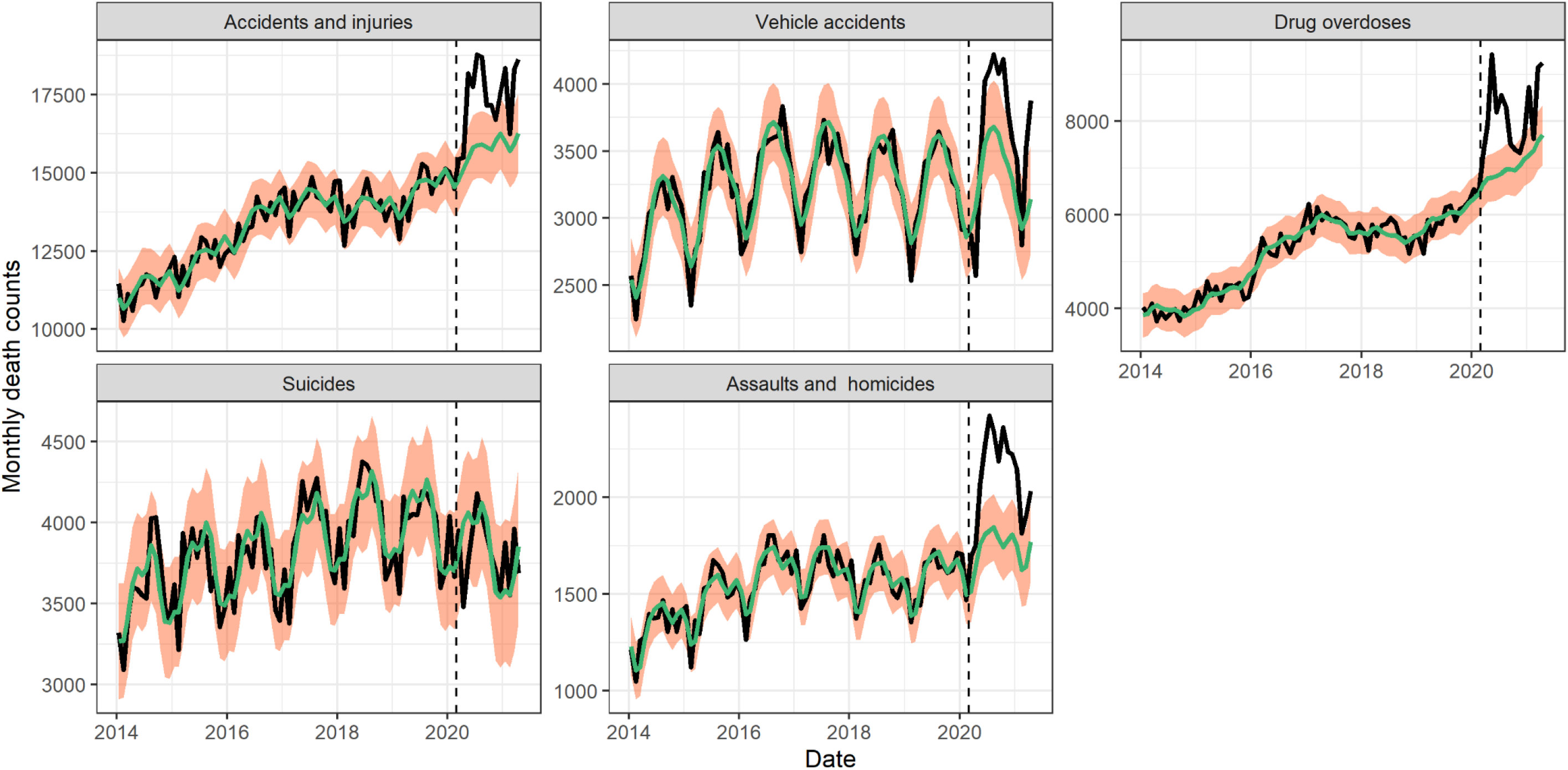
National monthly deaths by subcategory of external causes of death from January 1, 2014 to April 1, 2021. The black line shows observed data, the green line shows seasonal baseline, the orange shading represents the 95% CI on the seasonal baseline. The dotted vertical line marks the start of the pandemic on March 1, 2020.

**Table 3:**
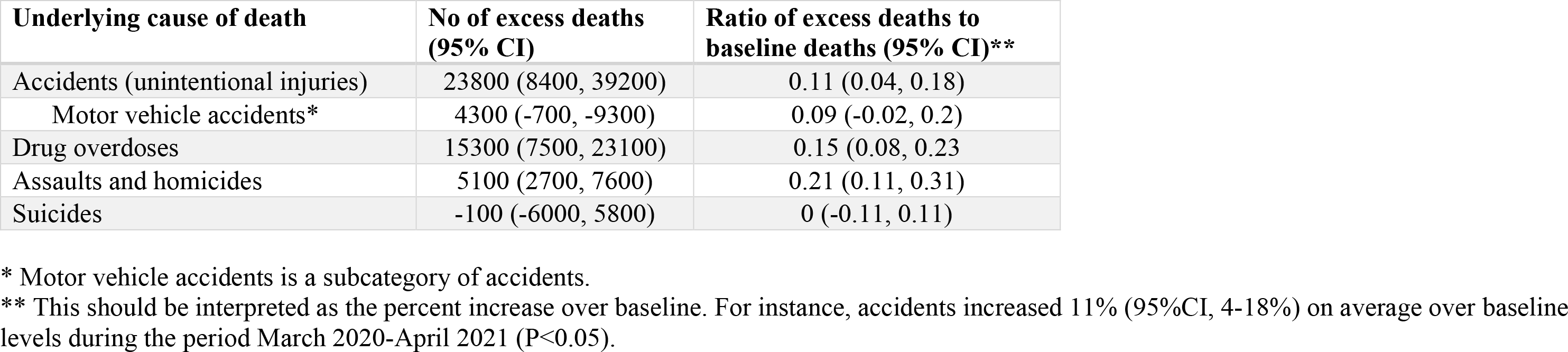
Excess mortality for different subcategories of external deaths during the COVID-19 pandemic period, March 2020 to April 2021.

We saw evidence of increased synchronicity in multiple causes of deaths during COVID-19, which is a possible signature of the direct effects of SARS-CoV-2 infection on mortality from chronic conditions. During the period March 2020-April 2021, all-cause mortality became more correlated with excess deaths from underlying respiratory conditions as compared to historical patterns in 14 out of 16 states (Figure S12). States that experienced high cumulative excess respiratory deaths had concomitantly high excess mortality from all-causes (Spearman rho = 0.73, 95% CI: 0.44 - 0.90)), attesting to the large impact of COVID-19 on total mortality (Figure S13). Synchrony between excess deaths from underlying respiratory diseases and excess deaths from underlying chronic conditions increased during the pandemic in a subset of states (Figure S12), particularly for diabetes (n=8 states), heart diseases (n=5), cerebrovascular diseases and Alzheimer (n=4). Excess deaths recorded as cancer, and external causes of deaths, showed no change, or declining synchrony, with respiratory mortality during the pandemic.

Next, to quantify the direct and indirect impacts of the pandemic on different causes of death, we regressed weekly cause-specific excess mortality against COVID-19 intensity and the strength of non-pharmaceutical interventions. On a national level, 90% (95% CI, 82-99) of excess deaths from all-causes were directly attributable to COVID-19, while the proportion was 59% (95% CI, 42 – 76) for diabetes, 58% (95% CI: 30 - 86) for Alzheimer’s, and 97% (95% CI: 40-158%) for heart diseases (Table 2). The upper bound of the 95% confidence interval for heart diseases was above 100% (158%), suggesting that for every excess death from heart disease estimated by our model, up to 1.58 death from heart disease could be directly linked to SARS-CoV-2 infection.

We also found evidence for the indirect effects of the pandemic on external causes of deaths and cancer. Periods of more stringent interventions were statistically associated with elevated mortality from external causes, while stricter interventions coincided with a decline in cancer mortality (Table S1). Analyses of direct and indirect effects at the state level yielded similar findings, with COVID-19 explaining 76 % (95% CI, 32-100) of all-cause excess deaths (Table S2). We did not identify predictors of cause-specific mortality at the state level, possibly due to lack of power (Table S2).

### Direct and indirect pandemic impacts by age group

The total burden and direct impacts of COVID-19 varied substantially by age (Figure 4, Table 4). Individuals 85 years and older had 155,100 (117300, 192100) excess all-cause deaths, accounting for 23% of excess deaths during the pandemic period. In contrast, individuals under 25 years and 25-44 years accounted for only 1.5% and 8.1% of pandemic excess mortality, representing 10,200 (6000, 14300) and 54,500 (47500, 61300) excess all-cause deaths.

**Figure 4.**
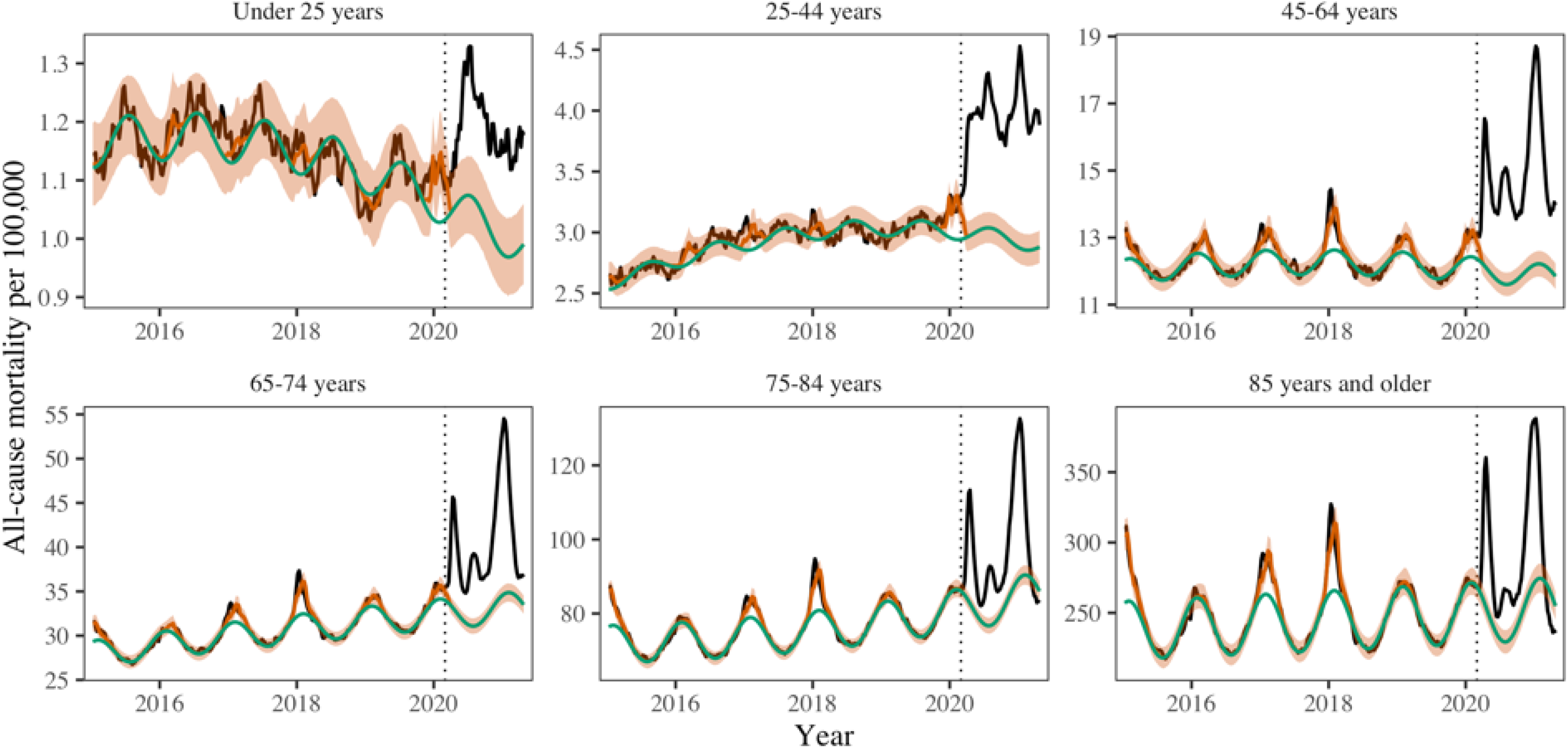
Age-specific all-cause mortality time-series graphs. The black line shows observed data, the green line shows seasonal baseline, the orange shading the 95% CI on the seasonal baseline, and the red line shows model predictions with seasonal variation and influenza circulation. The dotted red lines show the upper and lower 95% confidence intervals. Excess mortality attributed to the COVID-19 pandemic is defined as the area between the black and green line from March 1, 2020. The dotted vertical line marks the start of the pandemic on March 1, 2020.

**Table 4.**
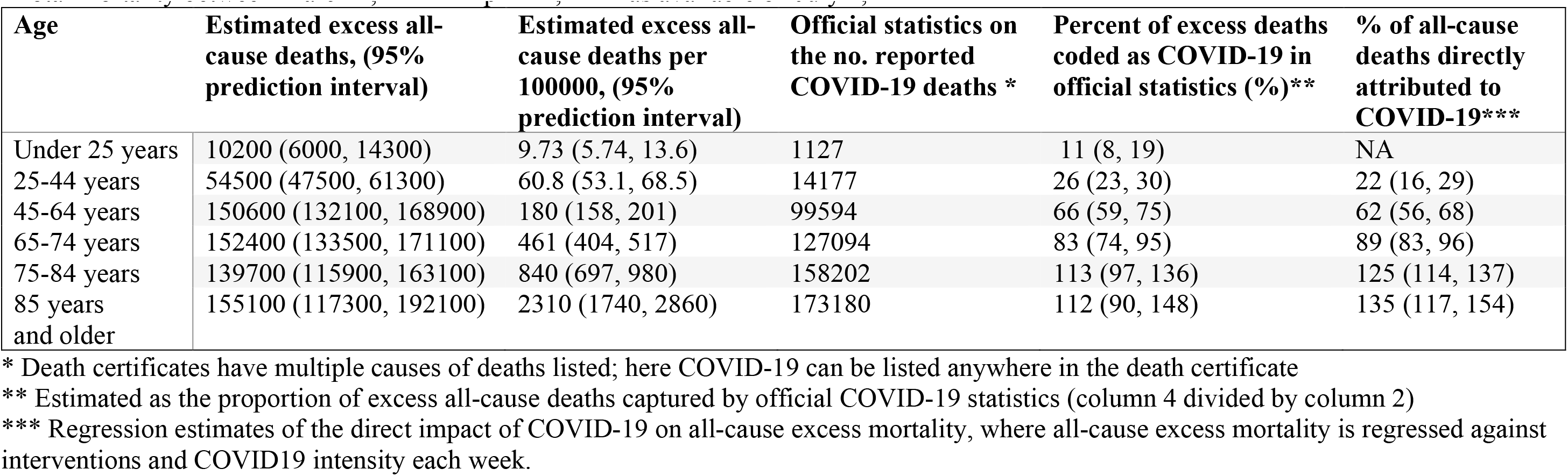
Excess all-cause deaths, official COVID-19 deaths, and direct contribution of COVID-19 to mortality, by age group. *Total mortality between March 1, 2020 – April 30, 2021 as available on July 5, 2021

Official COVID-19 statistics captured an increasing percentage of excess deaths with age, ranging from 11% (8-19%) in individuals under 25 years to over 100% in those over 85 years (Table 4). This age gradient is consistent with a larger direct effect of the pandemic in older age groups. We then used weekly regression of excess mortality on COVID19 activity and the strength of interventions to confirm these findings using a different approach and disentangle the direct and indirect pandemic effects. The COVID-19 term measuring the direct impact of viral infection was highest in older age groups and statistically significant in all age groups but those under 25 years. In contrast, the indirect impact of the pandemic measured by the intervention term was highest in youngest age groups, decreased with age, and lost significance in individuals above 65 years. This analysis indicates that deaths in younger ages rose in periods of stricter interventions, independently from the effect of SARS-CoV-2 circulation (Table S3). Finally, to better understand the interplay between indirect mortality in younger age groups and deaths from external causes, we visualized age-specific monthly statistics on external deaths. Excess deaths from external causes were concentrated in ages 15-44 years, with a notable elevation in May-July 2020 compared to historic baselines (Figure S14).

## Discussion

In this US study, we aim to disentangle the direct and indirect mortality impacts of the COVID-19 pandemic from March 2020-April 2021 using regression and synchronicity analyses. We find that 90% of the rise in all-cause mortality during this period can be statistically linked to SARS-CoV-2 infection, lending support to the predominance of the direct mortality consequences of the pandemic on a national scale. We also find a direct contribution of SARS-CoV-2 infections on chronic conditions such as Alzheimer’s, diabetes, cerebrovascular diseases, and heart diseases, which was missed in official statistics. Yet, analysis of mortality in children and young adults, and mortality from accidents and injuries, drug overdoses, assaults and homicides, paints a different picture. In these death categories, the mortality elevation observed during the pandemic period is statistically linked with interventions, supporting indirect pandemic effects rather than the direct consequences of SARS-CoV-2 infection. In contrast to other causes of deaths studied, cancer and suicides remained within baseline levels during the pandemic.

Perhaps the most striking finding of our study is the large mortality burden of the pandemic in individuals 25-44 years, with an estimated 54,500 (47,500, 61,300) excess deaths by April 2021. Only a quarter of these excess deaths are captured by official tallies of COVID-19 deaths. Accordingly, our regression analysis indicates that the majority of excess deaths in this age group are attributable to the indirect consequences of the pandemic. The trajectory of mortality in this age group is disjoint from the periods of intense COVID-19 circulation and statistically tied to the strength of interventions, supporting a possible detrimental effect of COVID-19 control measures beyond the initial lockdown period in Spring 2020. And even though children had a low absolute rate of excess mortality during the pandemic, only 11% was attributable to SARS-CoV-2 infection, while stricter interventions were associated with rising mortality in this age group. In contrast, we find that the predominant mortality impact of the pandemic in individuals over 65 years was associated with the direct consequences of SARS-CoV-2 infection. In a study of excess mortality in over 100 countries, Karlinsky and Kobak note that the direct mortality consequences of the pandemic typically predominate over the indirect consequences (Karlinsky & Kobak, 2021); however, they did not study age patterns. Our results support that the balance between direct and indirect effects is age-dependent and can skew towards indirect effects in the young, even in countries that experienced relatively high infections rates like the US.

Prior studies have shown that a decrease in emergency visits for diabetes, stroke, and myocardial infarctions in all age groups coincided with a rise in mortality for these conditions (Lange, 2020). Faust et al. estimated that 38% of deaths between 25 – 44 years were due to COVID-19 during March-July 2020, compared to 26% (23-30%) in our study that considers a much longer time period (Faust et al., 2020). Public health interventions, limited medical care, and behavioral changes (e.g., delays in seeking timely medical help due to fear of infection) (Bollmann et al., 2020; Kansagra et al., 2020; Mafham et al., 2020; Woolf et al., 2021) could have contributed to the surge in excess deaths unrelated to COVID-19 in young adults, resulting in a notable peak of mortality in summer 2020. In addition, we find that mortality from external causes remained elevated during May-August 2020 among young adults, likely driven by elevation in deaths from opioids, accidents, and assaults.

Mortality from external causes increased by 51,300 deaths (39700, 62700) from March 2020 - April 2021. There was a moderate correlation between excess mortality and pre-pandemic baseline rates for these causes (Figure S11), indicating that states with historically high death rates for suicide, drug overdose, and homicides experienced more prominent increases during the pandemic. The rise in mortality was most pronounced in assaults and homicide (21% over baseline, averaged over the pandemic period), followed by overdoses (15%) and accidents (11%). In contrast, mortality from suicides remained stable or below expectations. Prior work by Faust et al reported a decrease in suicide in several countries during March-July 2020, including in the US (10%) (Faust, Du, et al., 2021); here we show that suicide mortality remained stable throughout the rest of 2020. Further, Faust et al reported that deaths from overdoses and injuries increased during March-July 2020 (Faust, Du, et al., 2021). This is confirmed in our data, but we also show that the increase persisted in late 2020 and until the end of our study period in April 2021. Overall, of the 8 causes of death studied here, the indirect pandemic effects were statistically largest on external mortality causes.

Synchrony between respiratory mortality and other mortality causes was high during spring 2020, possibly due to poor SARS-CoV-2 test availability and guidelines to limit testing to just hospitalized cases. Many deaths in nursing homes or at-home during March-April 2020 were never tested, and they were recorded as known underlying conditions (i.e. heart disease, Alzheimer’s and diabetes) by default. In addition, Alzheimer patients typically live in nursing homes and may have been at increased risk of (untested) COVID-19 infection early in the pandemic. Interestingly, the correlation between excess mortality from respiratory diseases and Alzheimer increased in the winter 2020-2021 wave, signaling a persistent direct impact of COVID-19 on Alzheimer in a period where COVID-19 incidence and testing propensity were high.

We validated our excess mortality estimates against serology and assessed the IFR, a parameter notoriously difficult to measure. Our all-age estimate of 0.61% (0.49 - 0.73) is consistent with a recent meta-analysis (Meyerowitz-Katz & Merone, 2020) and aligns with an early study from China (0.66%; 95% CI: 0.39—1.33%) (Verity et al., 2020). A study of all-cause excess mortality in the Netherlands reports a substantially higher IFR (1%) (Asten et al., 2021); however, all-cause mortality is not specific to COVID-19 (our estimate based on all-cause mortality is higher at 0.86% (0.72 - 0.99)). IFR estimates based on official COVID-19 statistics were 15% higher than those based on excess respiratory mortality, yet the official statistics did not correlate with serology data as well as with our excess mortality estimates. Differences in COVID-19 death attribution by state could explain these findings. Overall, these analyses support the robustness of excess respiratory mortality as an indicator of COVID-19.

New York exhibited a notably high IFR. Several non-mutually exclusive factors could drive these findings, including a higher proportion of deaths among older individuals (aligned with the demography of New York state), large outbreaks in long-term care facilities, and lack of knowledge on management of severe patients early in the pandemic (Barnett et al., 2020). It is also worth noting that serosurveys conducted in April 2021 could underestimate cumulative SARS-CoV-2 infection rates in states that have experienced most of their infections in early 2020. Seroprevalence for New York state decreased from a peak of 23% in August 2020 to 13% in April 2021 in CDC data (Centers for Disease Control and Prevention, 2021f), which could be due to declining antibody titers (Buss et al., 2021) or sampling issues. To address this issue, we ran a sensitivity analysis using the maximum seroprevalence recorded over the study period rather than the seroprevalence at the end of the study period (Figure S9); New York remained an outlier in this analysis. Higher IFRs may be associated with healthcare systems working above capacity, as would have been the case in the first phase of the pandemic in New York.

The roll-out of a large SARS-CoV-2 vaccination campaign starting in late December 2020 in the US has had a major impact on rates of hospitalizations and deaths for COVID-19 in 2021. Accordingly, mortality from all causes and respiratory diseases declined continuously in the first months of 2021 and had just returned to baseline by the end of the study period in April 2021, before the arrival of new SARS-CoV-2 variants such as Delta and Omicron. It is interesting that at this particular point in the pandemic, in April 2021, mortality from cancer, Alzheimer and heart diseases was below baseline (negative excess mortality), with a similar phenomenon observed for all-cause mortality among individuals 75-84 and over 85 yrs. It is unlikely to be an artifact of reporting lags, as a reporting adjustment is built in the model, and reporting is 99% complete after 4 months. However, these negative excesses could signal a displacement of the mortality baseline, whereby frail individuals are harvested by large-scale infectious disease events or heatwaves, resulting in a decline in baseline mortality in the aftermath of an acute mortality event (Saha et al., 2013). Harvesting is also consistent with our regression analysis, where estimates of the direct impacts of COVID19 exceeded 100% in the two oldest age groups (albeit with broad confidence intervals, Table 4). This would be expected if baseline mortality was overestimated due to harvesting. Further, the age profile of COVID-19 severity risk dictates that older individuals would bear the strongest effects of harvesting, which is consistent with the US data. We can’t rule out the putative effect of SARS-CoV-2 vaccination on reducing baseline (non-COVID-19) mortality, although a direct biological mechanism is difficult to pinpoint.

Our study is subject to several limitations. First, mortality counts below the minimum cut-off value of 10 were suppressed due to privacy regulations. As a result, our age-specific analyses are restricted to larger states, and we could not assess the role of race/ethnicity. Prior work has shown important disparities in COVID-19 impact by race/ethnicity and economic status (Mena et al., 2021; Rossen, 2021) in the US and abroad. Second, official coding practices may have changed between states and through time based on SARS-CoV-2 testing availability, location of death, demographic factors, and comorbidities. Third, we find periods of negative excesses in cancer (throughout the pandemic), cardiovascular, and heart diseases (fall 2020), possibly due to changes in ascertainment of underlying cause of death (e.g. a death in a cancer patient with COVID-19 is ascribed to COVID-19) or harvesting (Saha et al., 2013). As discussed earlier, harvesting could also have affected estimates in oldest age groups. Similarly, we can’t account for changes in baseline respiratory mortality due to depressed circulation of endemic pathogens other than influenza. Finally, our study ends in April 2021 and does not capture a recrudescence of COVID19-related deaths due to the more transmissible Delta variant, primarily in states with low vaccine coverage, nor do we estimate the impact of the Omicron immune escape variant. As a result, our excess mortality estimates should be deemed conservative.

Pandemic excess mortality patterns have been heterogeneous globally (Islam et al., 2021; Karlinsky & Kobak, 2021; Kontis et al., 2020; Nørgaard et al., 2021). In a comprehensive analysis of mortality in 21 countries in Europe, New Zealand and Australia, official COVID-19 deaths accounted for an average of 77% (62%-93%) of all-cause excess deaths during the first wave of the pandemic March-May 2020 (Kontis et al., 2020). However, there was greater disconnect between estimates in hard-hit countries such as the UK, Spain, Italy, and Belgium (Kontis et al., 2020; Kontopantelis et al., 2021; Odone et al., 2021). In a study of over 100 countries, the ratio between excess mortality and official deaths was 1.6 on average, but went as high as 50 (e.g. Nicaragua) (Karlinsky & Kobak, 2021). The disconnect was primarily attributed to under-detection of COVID-19 rather than indirect effects, although indirect effects were not explicitly modeled. In Italy, the case fatality rate for acute myocardial infarction increased 3-fold during the first wave, while hospitalizations for these conditions decreased by 48% (Odone et al., 2021), suggesting that at least some of the excess deaths were indirect deaths. In the UK, cancer deaths increased about 10% at the height of the April 2020 lockdown; however, more recent fluctuations in cancer mortality remain unclear (Lai et al., 2020). Interestingly, in New Zealand, where control of COVID-19 has been remarkable, mortality was slightly but not significantly below baseline (Kontis et al., 2020). A similar finding was described in Russian provinces where a lockdown was implemented before the onset of COVID-19 (Kobak, 2021). This suggests that a lockdown without COVID-19 is neither preventing nor causing an appreciable number of deaths, although effects could be country-dependent. In the US, the direct impacts of the pandemic greatly outweigh the indirect consequences in all age data, but the reverse is true in children and young adults. Further work should concentrate on comparing the direct and indirect impact of COVID-19 in different countries over the same time period and using the same methodology.

## Conclusion

Here, we examined trends in cause-specific mortality across states and age groups to address the direct and indirect impacts of COVID-19 in the US. We find that ∼90% of the total mortality elevation during the March 2020-April 2021 pandemic period is attributable to the direct impact of SARS-CoV-2 infection. There is however a large indirect impact of the pandemic in children and young adults, and on mortality from external causes, particularly from accidents, assaults, and overdoses. The indirect mortality impact of the pandemic is statistically linked to the strength of interventions. We also find an undetected contribution of SARS-CoV-2 infections on mortality from chronic conditions, such Alzheimer’s, diabetes, cerebrovascular conditions, and heart diseases, that persisted throughout winter 2020-2021. Our conclusions are based on ecologic analyses that are useful to generate hypotheses but do not prove causality. As more detailed mortality information become available with the release of death certificate data, it will be important to dissect the drivers of mortality among younger adults and certain ethnic groups, and understand how chronic conditions, violence, opioids and suicides intersect with large-scale infectious disease events and behavioral changes.

## Data Availability

Data and code are posted on github

## Acknowledgements

DMW acknowledges funding from NIH/NIAID (R01AI137093). LS acknowledges funding from the Carlsberg foundation.

## A. Supplemental Data and Methods

### 1. General approach

Our analysis relies on modeling of weekly trends in US death certificates compiled from the National Center for Health Statistics (NCHS) website ^1^. The goal of the study is to estimate the direct mortality impact of COVID-19, which results from SARS-CoV-2 infection, from the indirect impacts of the pandemic which can be linked to societal or health-related changes brought about by the pandemic.

We define direct COVID-19 mortality as the sum of deaths confirmed and coded as COVID-19 as the underlying cause, deaths coded as another underlying primary cause but with COVID-19 as one of the contributing multiple causes, and deaths that were directly caused by SARS-CoV-2 infection but for which a COVID-19 code is not included in the death certificate due to misdiagnosis or lack of testing. The first two categories are included in official death tallies of COVID-19 (since a COVID-19 code appears in the death certificate), while the third category can only be estimated using excess mortality approaches.

We define the indirect mortality impact of the pandemic as the sum of deaths due to healthcare avoidance or inaccessibility, deaths due to conditions or events that are exacerbated by non-pharmaceutical interventions and other pandemic behavior, and would not have occurred otherwise (i.e. suicide, drug overdose, homicide, or a stressed healthcare system that is unable to treat conditions unrelated to SARS-CoV-2).

It is worth noting that circulation of multiple pathogens has plummeted due to social distancing interventions in 2020-2021,^2^ and therefore the pandemic could have prevented a number of infectious disease deaths relative to historical expectations. Excess mortality approaches^3,4^ will capture the net sum of these indirect impacts. These direct and indirect mortality pathways are not mutually exclusive and plausible over a wide range of conditions. For example, SARS-CoV-2 infection may trigger death in a patient with diabetes, which may be missed by testing, with no COVID-19 code listed on the death certificate. Concomitantly, a diabetic patient without a recent history of SARS-CoV-2 infection may turn away from the healthcare system at the height of the pandemic and die from lack of treatment. Our analysis attempts to separate these effects.

### 2. Mortality Data

#### a. Mortality conditions studied and states selected for further analysis

We used the international classification of disease version-10 to retrieve deaths for the following 8 specific conditions: All-cause (deaths from any causes), Alzheimer’s (G30), Cancer (C00-C97), Cerebrovascular diseases (I60-I69), Diabetes (E10 – E14), Heart disease (I00-I09, I11,I13,I20-I51), Respiratory Conditions (J09-J18, J40-J47, U071), External Cause (V01-Y89, U01-U03). Deaths with any of these codes as the <u>underlying</u> cause of deaths were selected for analysis.

16 states were selected for further analysis of respiratory mortality because they had sufficient counts on a weekly basis: Alabama, Arizona, California, Florida, Georgia, Illinois, Indiana, Michigan, Missouri, New Jersey, New York, Ohio, Pennsylvania, Tennessee, Texas, Virginia. Weekly deaths count below 10 are blanked by NCHS for privacy reasons; states that had more than 2 weeks of blanked observations during the pandemic period were excluded. Respiratory mortality was our most restrictive mortality outcome; it is based on aggregation of deaths from pneumonia and influenza with deaths from other respiratory conditions. Pneumonia and influenza can be uncommon on a weekly basis in small states, especially in summer, and hence blanked observations are not uncommon in the state-level dataset.

We applied a similar reasoning to the 7 other causes of deaths that are unrelated to respiratory conditions. Death counts were more numerous for these conditions than for respiratory mortality. If a state had more than 2 blanked weeks for one of the conditions, it was excluded for analysis of the other conditions. The following 33 states were included for analysis of non-respiratory mortality: Alabama, Arizona, Arkansas, California, Colorado, Connecticut, Florida, Georgia, Illinois, Indiana, Iowa, Kansas, Kentucky, Louisiana, Maryland, Massachusetts, Michigan, Minnesota, Mississippi, Missouri, Nevada, New Jersey, New York, Ohio, Oklahoma, Oregon, Pennsylvania, South Carolina, Tennessee, Texas, Virginia, Washington, Wisconsin.

### 3. Analytical approach

#### a. Adjustment for reporting delays

Death counts are not finalized until several weeks after occurrence in the NCHS database. To estimate reporting delays, and for consistency with a prior analysis ^3^, we used a modified version of the NobBS package in R. The algorithm uses “snapshots’’ of data downloaded across different periods to infer reporting delays and the completeness of data at each reporting interval, which are then used to estimate true death counts. For this study, we used data from the NCHS website^1^ downloaded every Friday for 25 weeks (July 17, 2020–January 22, 2021, with the exception of week 2020-10-02). This algorithm was used to adjust reporting delay for all-cause and cause-specific mortality, allowing reporting delays to vary by cause and state.

#### b. Weekly excess Mortality Model

We applied seasonal regression models to weekly cause- and age-specific mortality, inspired by prior work on COVID-19 ^3,4^. Models included time trends, harmonic terms for seasonality, and terms for influenza circulation, following:

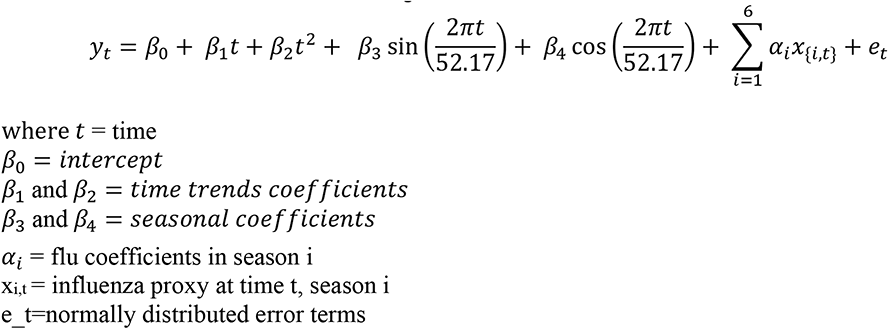

The proxy for weekly influenza incidences was calculated by multiplying the weekly percentage of physician visits for influenza-like illness and weekly percentage of positive influenza tests, ^4^ which were obtained via CDC’s FluView portal using the cdcfluview package in R (package version 0.9.1). We let the influenza coefficient vary each season to reflect a different mix of circulating subtypes, associated with different severities. Influenza data were not available for New Jersey and Florida; instead, we used data from New York state and HHS Region 4, respectively. Some states discontinue laboratory surveillance for influenza virus circulation during the summer months, so we replaced the missing summer weeks with zero. Weekly influenza incidences after March 1, 2020 were set to zero, in line with the minimal circulation of influenza reported in this time period. ^4^ Computation of confidence intervals aligns with prior work on COVID-19 excess mortality in the US ^3^.

March 1, 2020 was set as the start of the period of putative pandemic-related excess mortality. We fitted the model to data until March 1, 2020 and projected the baseline forward until April 30, 2021. We estimated weekly excess mortality related to the COVID-19 pandemic by subtracting the predicted baseline from the observed mortality that week. Total excess mortality for the pandemic period was defined as the sum of weekly excesses (positive or negative) during March 1 – April 30, 2021.

Of note, the impact of seasonal influenza was estimated by having an explicit flu coefficient in the excess respiratory model, while the COVID-19 impact was estimated by taking the difference between the observed mortality and baseline mortality during the pandemic period. Generally, this would be expected to inflate the impact of COVID-19, relative to influenza. But given that our COVID-19 estimates based on respiratory deaths align particularly well with official COVID-19 counts, and circulation of respiratory pathogens was minimal during this time period except for SARS-CoV-2^2^, we believe that our comparison of excess respiratory deaths between COVID-19 and seasonal influenza is fair.

We also tested different link functions and error structures for the model and report the best fit to data here.

#### c. Monthly excess mortality model for subcategories of external deaths

To better understand the rise in external deaths during the pandemic, we applied a similar excess mortality approach to deaths from 5 subcategories of external deaths available at a monthly resolution^5,6^: all accidents, motor vehicle accidents, drug overdoses, assaults and homicides and suicides. Because these data were on a monthly time scale, and less stationary that the other mortality causes, we tried more flexible model formulations including spline terms for time trends and/or for seasonality. Based on AIC, a model with spline terms for time trends and seasonality provided the best fit to the data. We did not include a term for influenza as there is no biological reason for why influenza would affect external deaths. External cause of death data were not available by subcategory and age, but we had age-specific data for all external causes of death combined ^6^. We visually explored these age-specific data as there was not enough information to fit time series models (Fig S14).

#### d. Estimation of the direct and indirect mortality impacts of the pandemic

We use a two-step approach to estimate the direct and indirect impacts of the pandemic. In the first step, we estimate the national weekly excess mortality for a given cause of death and age group, along with confidence intervals. In a second step, we regress weekly cause- and age-specific excess mortality against the weekly strength of non-pharmaceutical interventions (Oxford Health containment index^7^) and weekly COVID-19 activity (proxied by the weekly number of official tallies of COVID-19 deaths in NCHS data). The number of excess deaths from any cause directly attributable to COVID-19 is given by the regression coefficient for COVID-19 multiplied by the cumulative sum of the weekly COVID-19 predictor. A similar logic applies to the contribution of interventions on mortality. We explored potential delays between excess mortality and covariates (intervention measures and COVID-19 activity) using cross-correlation analysis (ccf function in R). We identified a lag of 4 weeks for interventions, and no lag for COVID-19, consistent across mortality outcomes and age groups.

To propagate the uncertainty in excess mortality estimates (response variable) between the first and second regression steps, we resampled the weekly excess mortality estimates based on their mean estimated values and standard deviation provided by the step 1 seasonal regression model, assuming a normal distribution. We sampled excess mortality 1000 times at each week (generating 1000 time series of excess mortality) and performed univariate and multivariate regression using COVID19 activity and interventions as covariates. Then, for each regression, we approximated the estimated slope coefficients and associated confidence intervals using normal distributions and sampled from those 100 times. This resulted in 100,000 slope estimates, from which we estimated the final confidence intervals for the effect of COVID19 and interventions on different age groups and causes. This method allowed us to account for the uncertainty in the response variable as well as the uncertainty in each fit.

We ran a similar approach to explore direct and indirect mortality effects in state-and cause-specific data. We regressed the cumulative excess mortality rate for the March 2020-April 2021 pandemic period for a given state and cause of death against average COVID19 official death rate, interventions, and cause-specific mortality baseline for the same period and state. The inclusion of a mortality baseline predictor was used to test the hypothesis that states that have high mortality for a given condition in typical years also fare worse during the pandemic. We propagated uncertainty in the response variable in the same way that we did for the weekly national estimates. For all states and causes of death, we ran univariate analyses and used AIC for variable selection in multivariate models.

Here we used a two-step approach to estimate direct and indirect mortality effects (Step 1: estimate weekly excess mortality and Step 2: regress weekly excess mortality against weekly COVID19 activity and weekly interventions). An alternative approach would be to run a single model, where weekly mortality rates are regressed against seasonal terms, time trends, flu activity, COVID-19 activity, and interventions. However, because there were only 60 pandemic weeks in our dataset, compared to 292 pre-pandemic weeks where the COVID19 and intervention coefficients are zero, that would be insufficient to get a robust estimate of the COVID-19 and intervention coefficients.

#### e. Estimation of the Infection Fatality Ratio

Without accounting for delays between infection and death, the infection fatality ratio (IFR) can be estimated by dividing the total number of deaths by the total number of infections. In other words, we have:

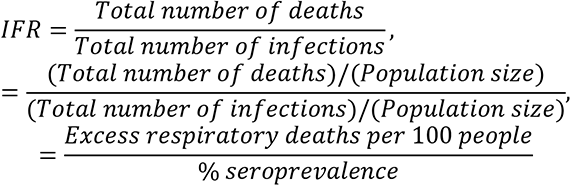

assuming that excess respiratory deaths provide a good estimate for deaths attributable to COVID-19. If we have information on excess deaths and serology in multiple states, we can regress these factors against each other, and the slope gives the average nationwide IFR when the intercept is set to zero (Figure 1). Given that the delay between seroconversion after infection is around 2 weeks, and the delay between infection and death is in the same order of magnitude, we use excess deaths until April 30^th^, 2021 in the numerator and serology for the last week of April 2021 in the denominator.

To propagate uncertainty from both the response variable (excess mortality) and covariate (seroprevalence) into IFR estimates, we used a similar approach as for the direct and indirect attribution model in the previous section. We resampled from the reported estimates of excess mortality and seroprevalence in each state, assuming normal distributions aligned with the reported 95% CI. Then, we drew 10,000 samples of excess mortality and seroprevalence in each state, and for each sample data set, we performed a linear regression and estimated the slope. Next, we drew 100 random samples for each of the 10,000 slope distributions. We calculated the confidence intervals of our IFR estimate by aggregating random samples of IFR estimates across all 100,000 sample data sets and taking 2.5% and 97.5% quantiles.

---

**Figure S1:**
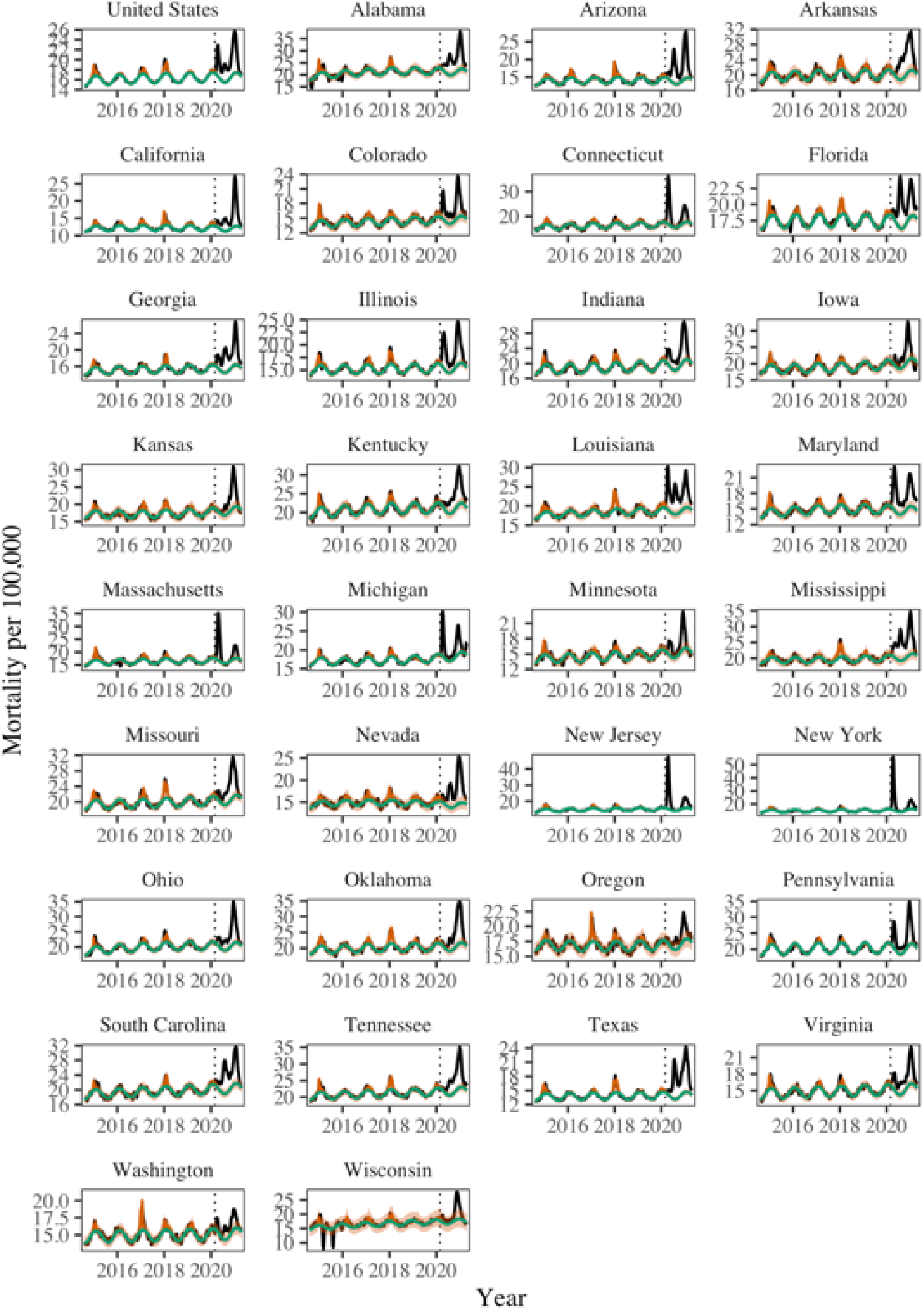
Trends in weekly all-cause mortality, nationally and by state. Black lines show observed data. Green line shows the seasonal baseline. The red solid line shows seasonal variation accounting for influenza circulation. The orange shading shows the upper and lower 95% confidence intervals. The dotted vertical line marks March 1, 2020.

**Figure S2:**
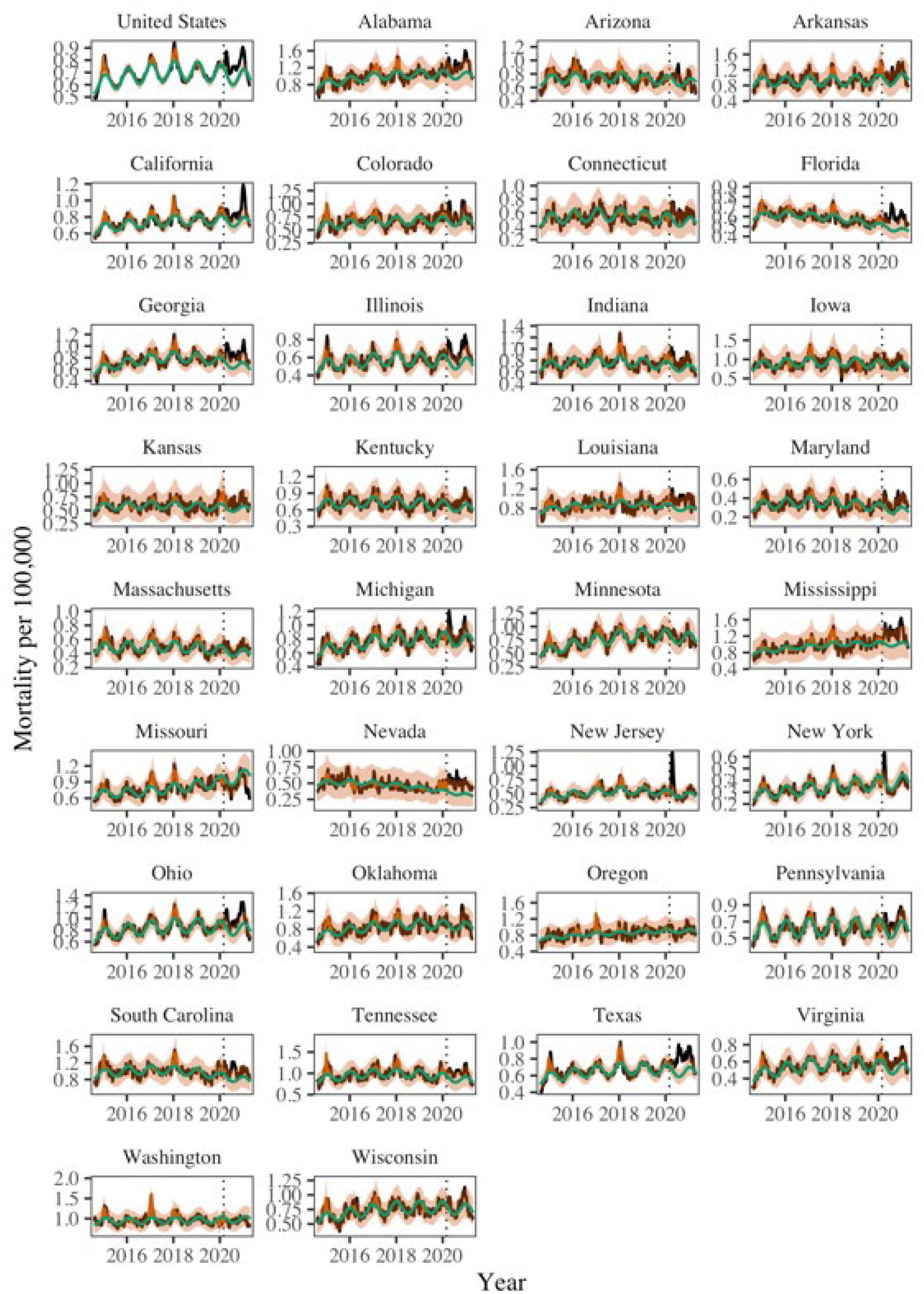
Trends in weekly Alzheimer mortality, nationally and by state. Legend as in Figure S1

**Figure S3:**
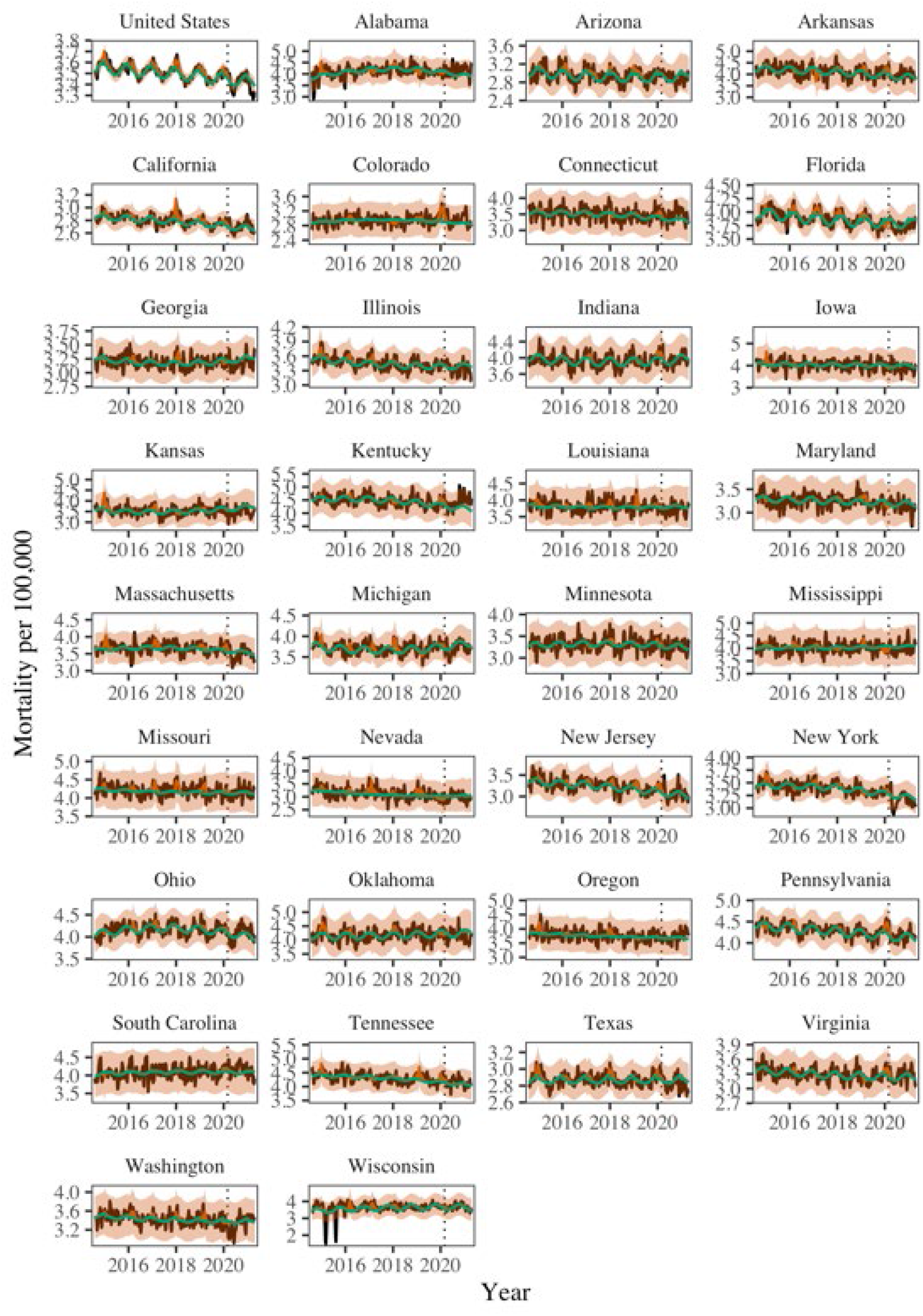
Trends in weekly cancer mortality, nationally and by state. Legend as in Figure S1

**Figure S4:**
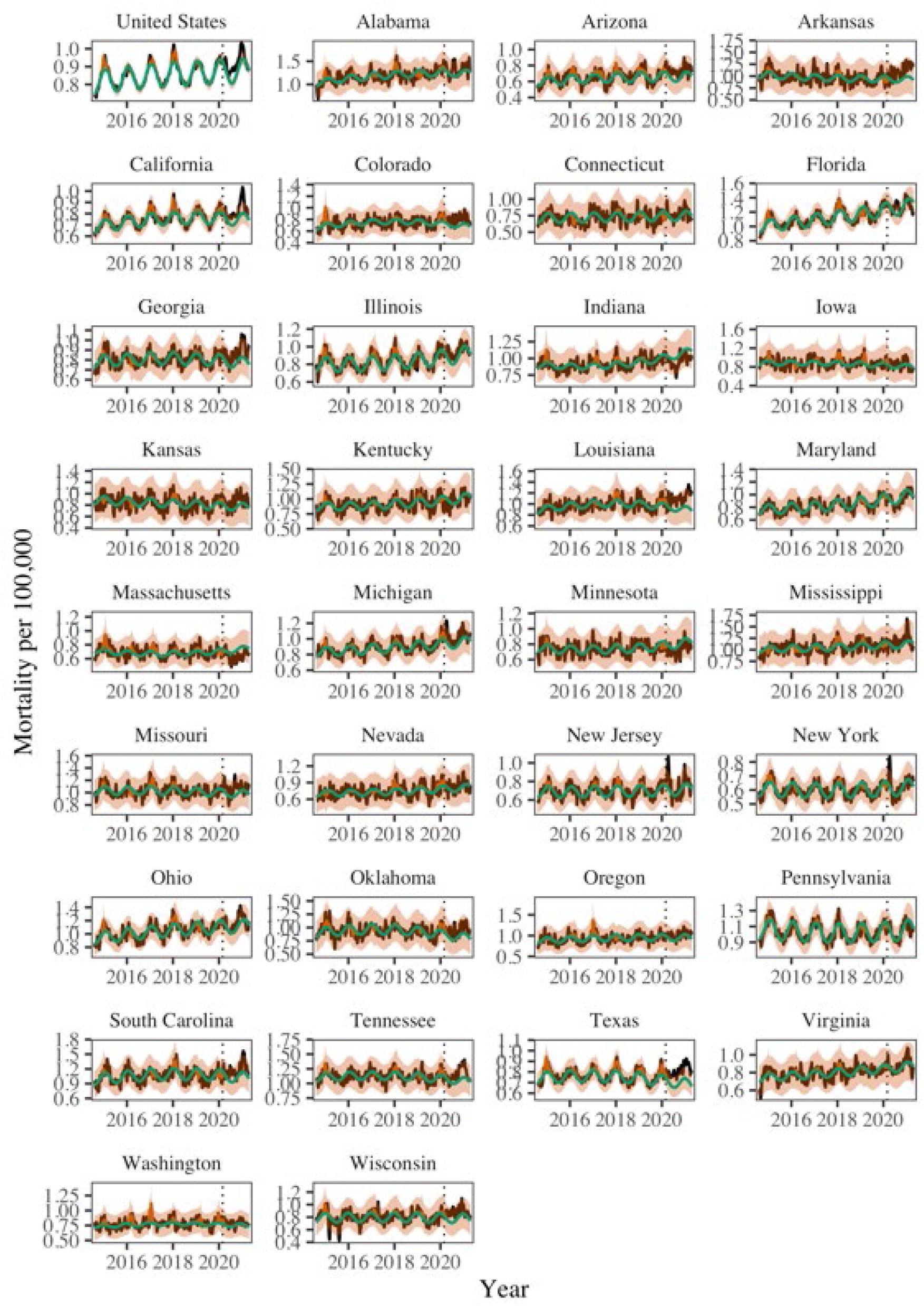
Trends in weekly cerebrovascular disease mortality, nationally and by state. Legend as in Figure S1

**Figure S5:**
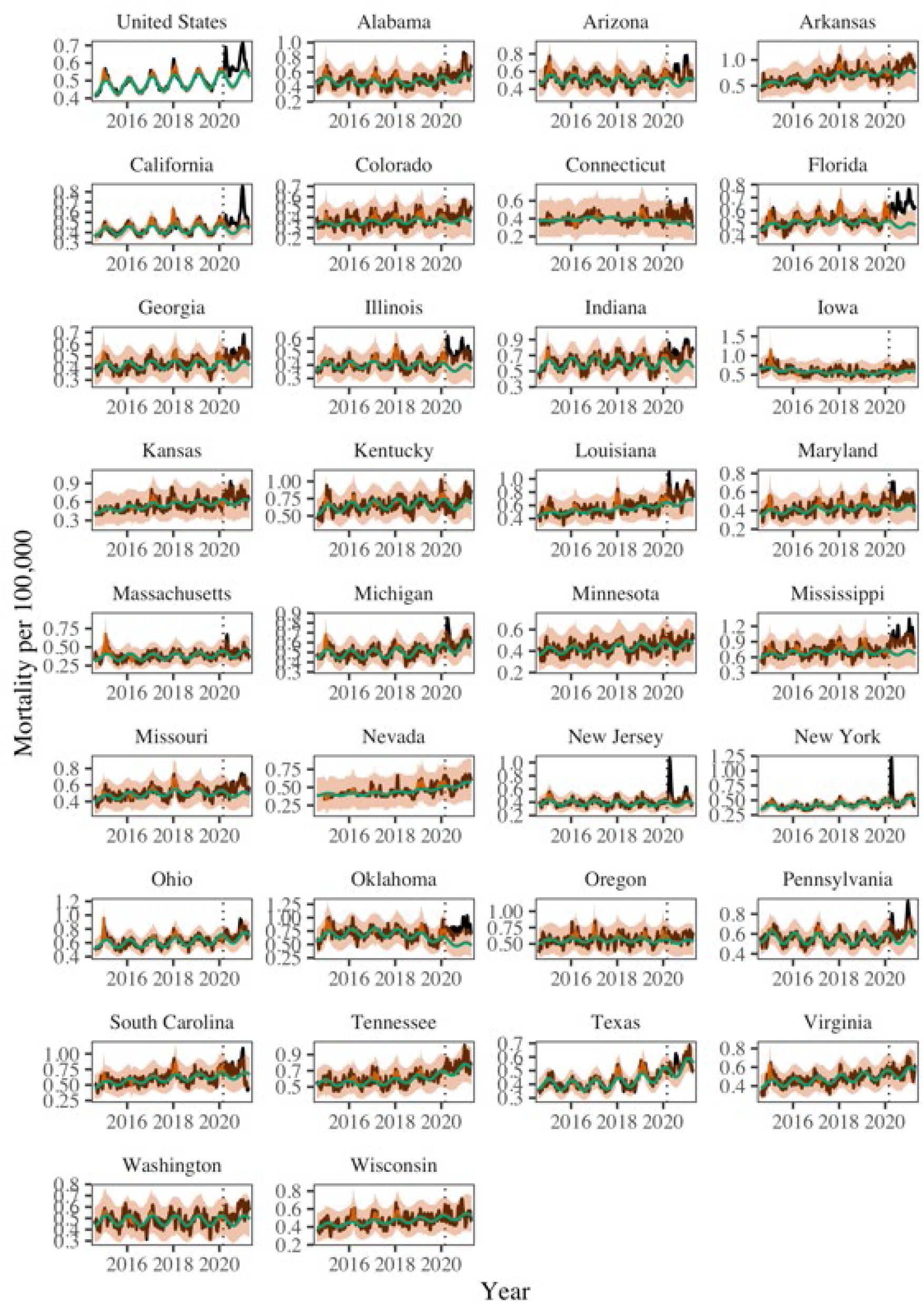
Trends in weekly diabetes mortality, nationally and by state. Legend as in Figure S1

**Figure S6:**
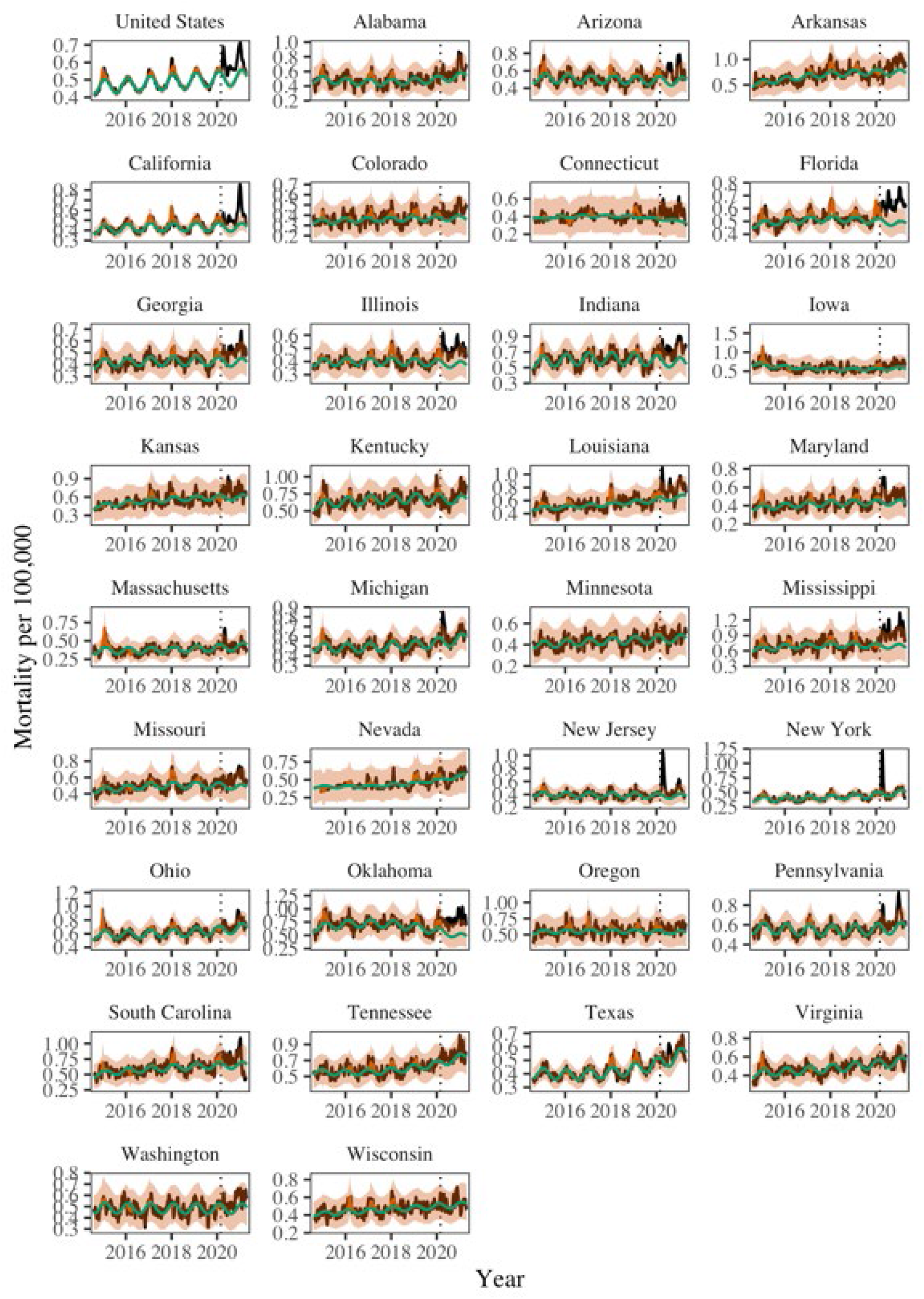
Trends in weekly heart disease mortality, nationally and by state. Legend as in Figure S1

**Figure S7:**
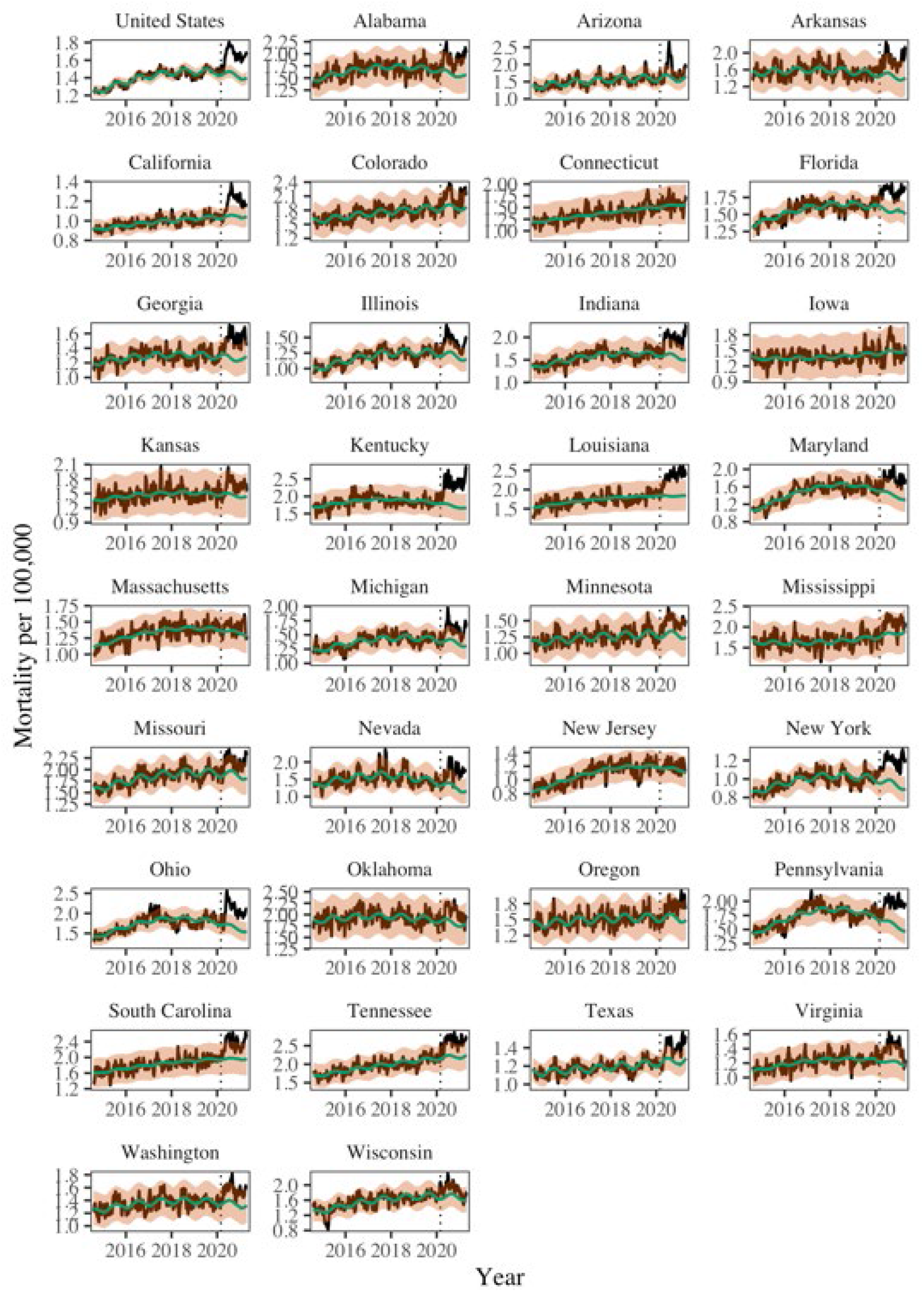
Trends in weekly mortality from external causes (opioids, suicides, accidents, etc.), nationally and by state. Black lines show observed data. Green line shows the seasonal baseline and the red shaded region shows the upper and lower 95% confidence intervals. The dotted vertical line marks March 1, 2020.

**Figure S8:**
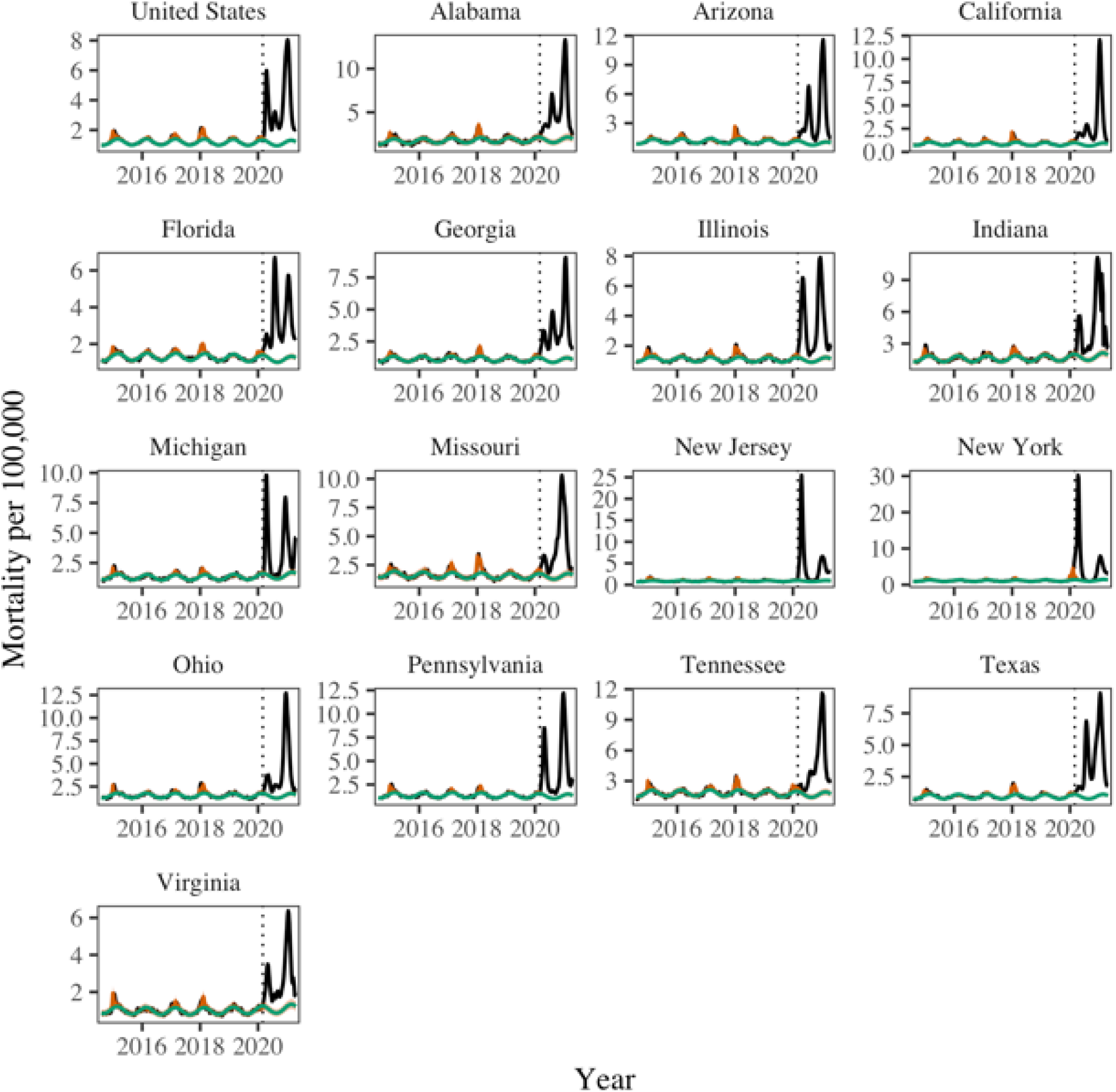
Trends in weekly respiratory mortality, nationally and by state. Legend as in Figure S1 (only 16 states had sufficient weekly data).

**Figure S9:**
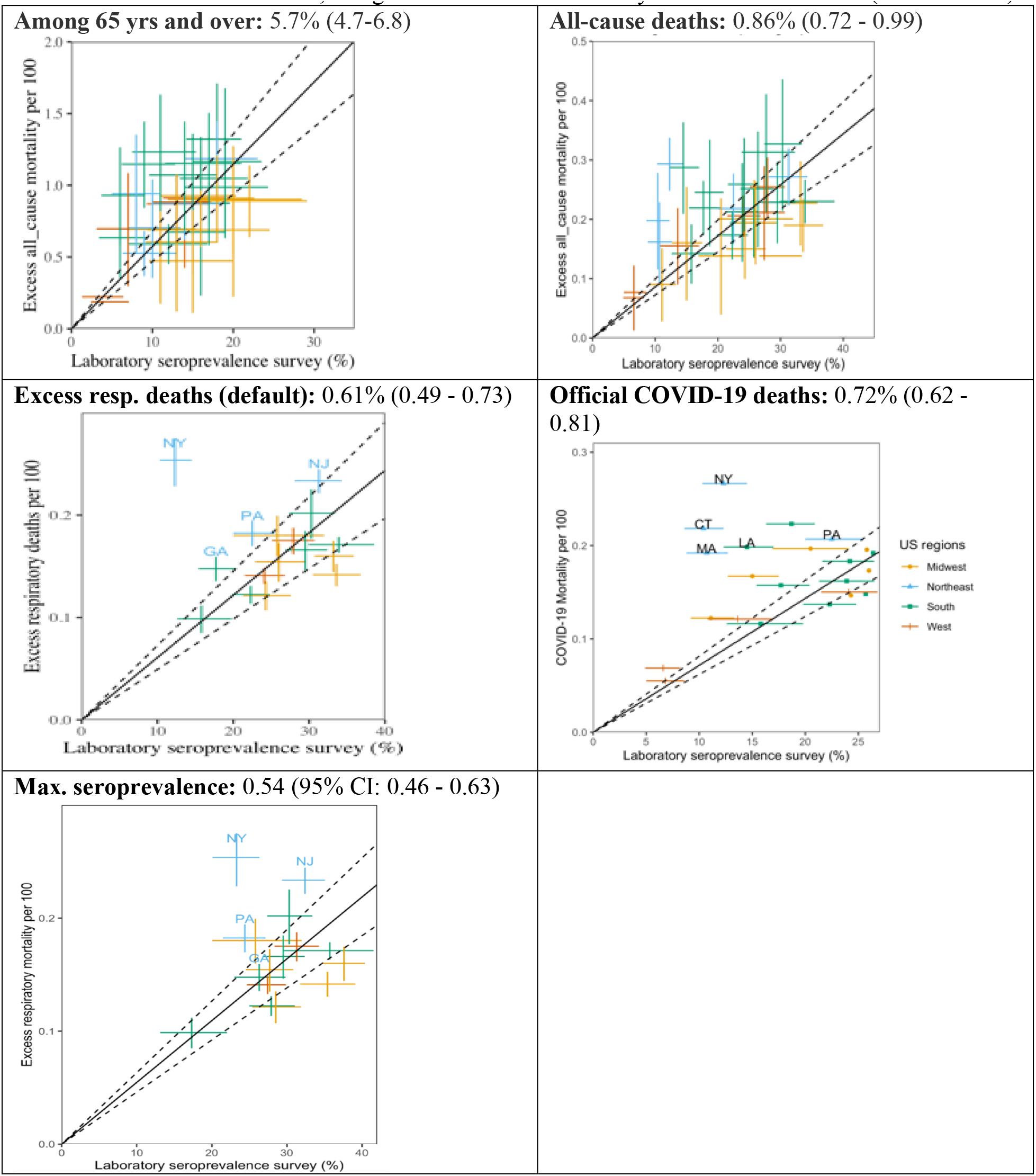
Sensitivity Analyses of Infection Fatality Rate (deaths/infections). In sensitivity analyses, we considered different indicators of COVID19 mortality in numerator (excess respiratory deaths, excess all-cause deaths or official COVID-19 death counts), and infection estimates based on seroprevalence in denominator (seroprevalence at the end of the study period, or maximum over the study period), and different age groups (all ages or over 65 yrs, both in numerator and denominator). COVID-19 seroprevalence estimates are from the last week of April 2021. Each point corresponds to a state. States with particularly high infection fatality ratios are annotated. Error bars represent 95% confidence intervals. The black line and dotted region represent a linear regression fit and the associated 95% confidence interval. For instance, using official COVID19 tallies yields an IFR of 0.72% (0.62 – 0.81%).

**Figure S10:**
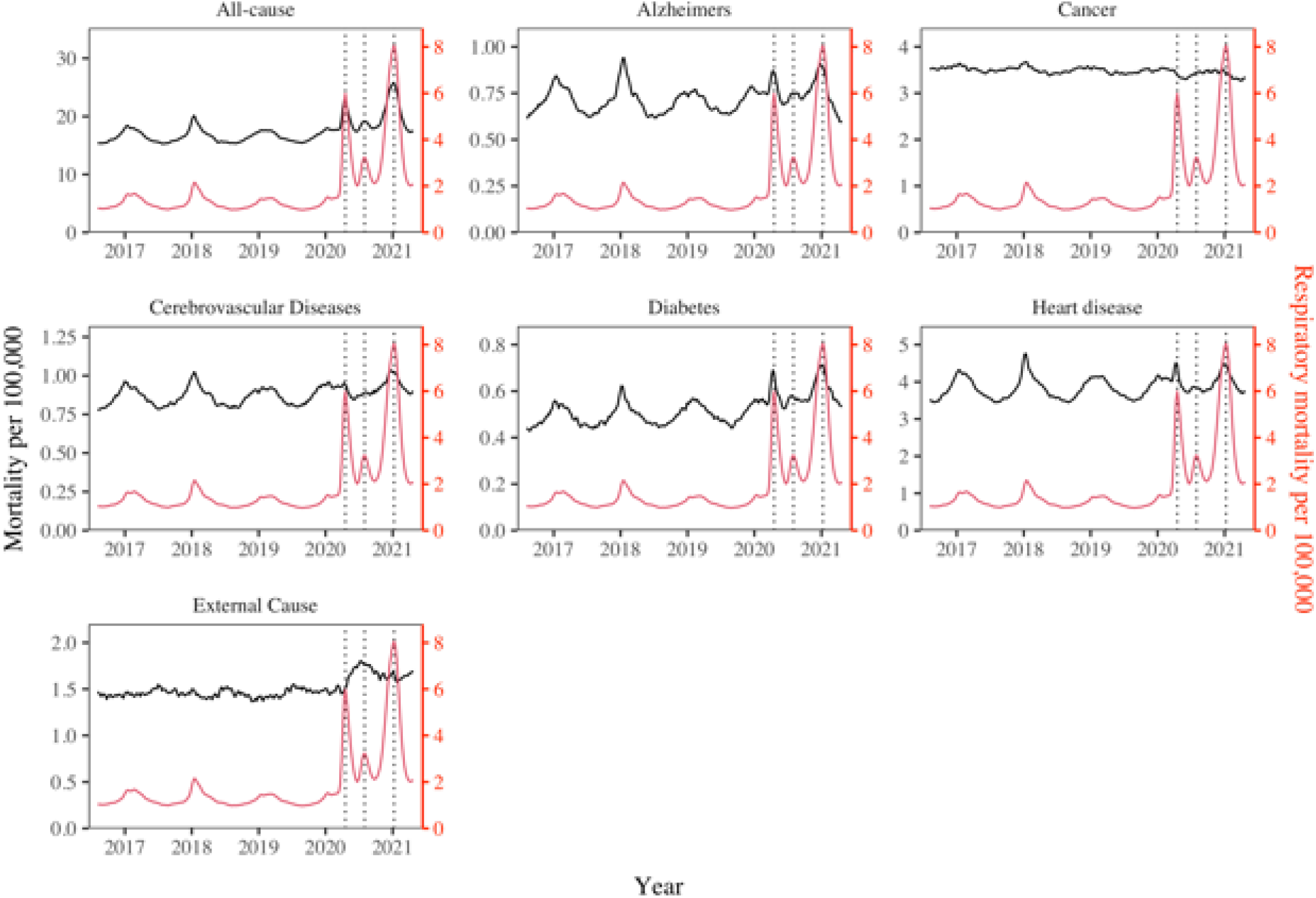
Synchrony between respiratory and non-respiratory mortality patterns on a national scale. The black lines show the time-series for each non-respiratory mortality cause. The red line is respiratory mortality. The dotted black lines mark the dates of the peaks during the first, second, and third wave in respiratory mortality (April 18, 2020; August 1, 2020; January 9, 2021).

**Figure S11:**
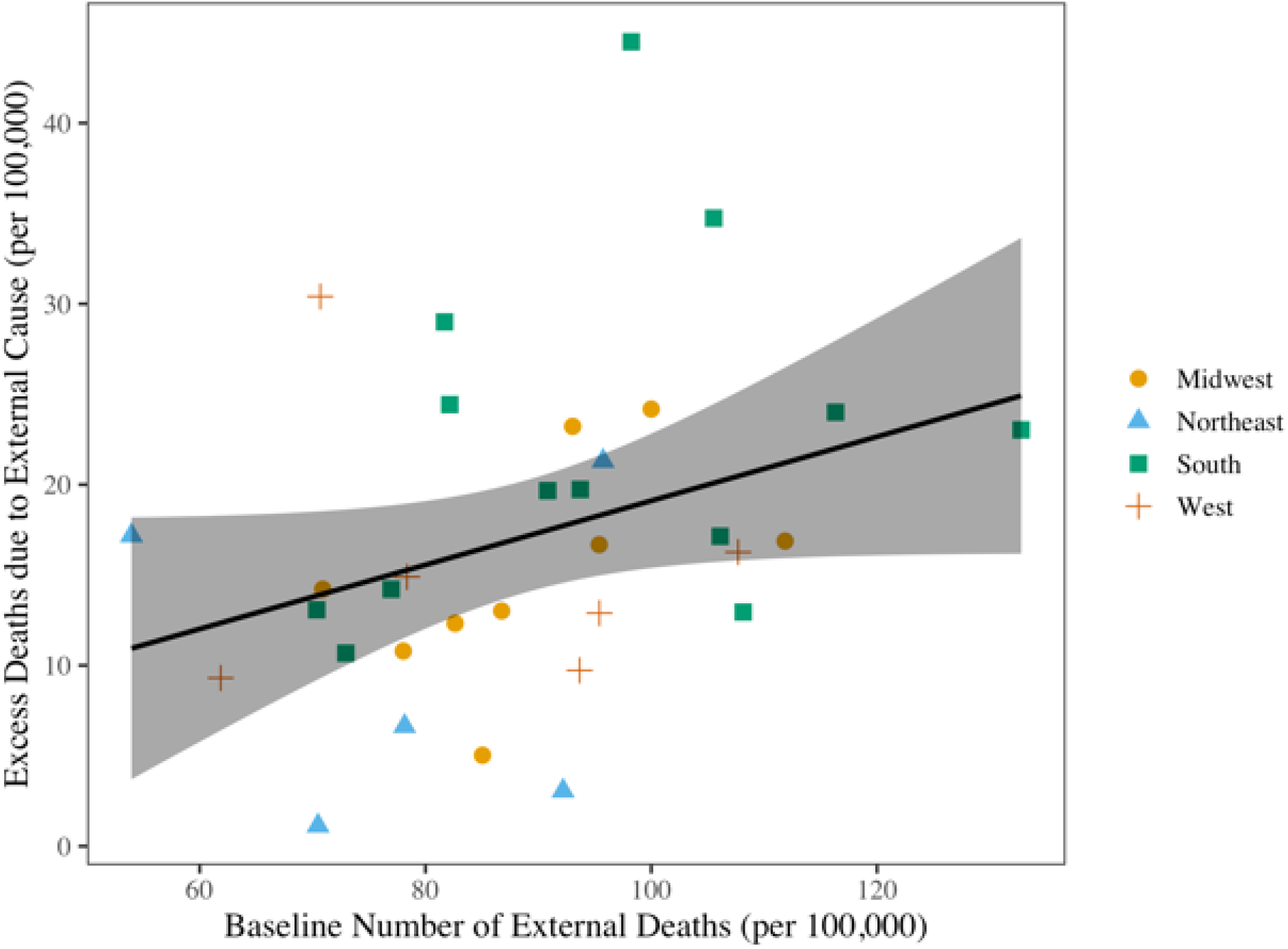
Correlation between cumulative excess death rates due to external causes (opioids, suicide, accidents, etc.) during March 2020-April 2021 and baseline death rates of external causes, across 33 states. A moderate correlation in these data indicates that states that typically have high rates of mortality from external causes fared worse during the pandemic.

**Figure S12.**
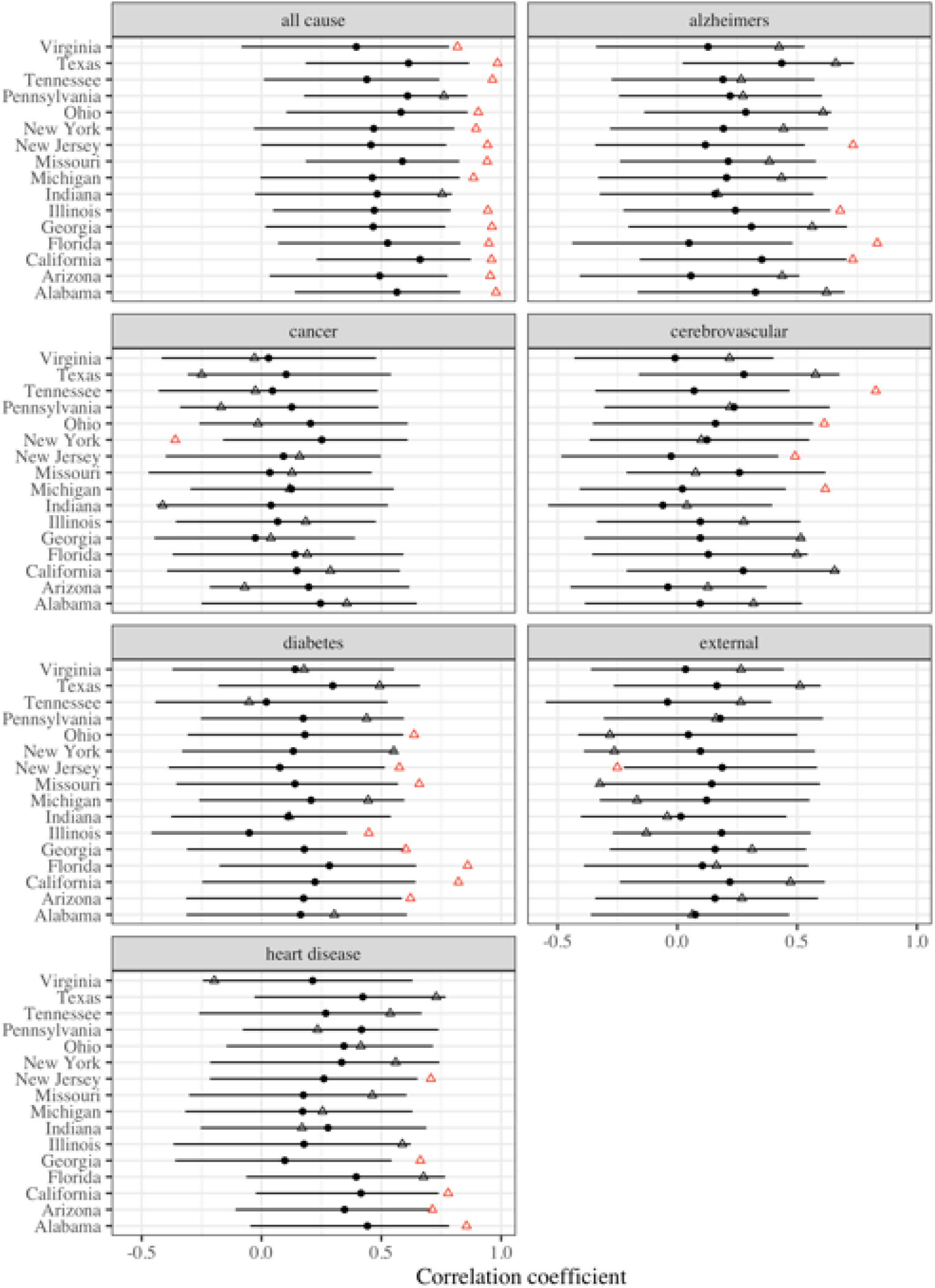
Changes in weekly synchrony between respiratory and non-respiratory mortality during the pandemic. Graph compares correlations during 60 weeks of any baseline pre-pandemic period and in the 60 weeks of the pandemic. Black points represent estimated pre-pandemic correlations (60 weeks selected before March 2020 by block of 2 weeks). Black error bars represent 95% bootstrap confidence intervals accounting for multiple comparisons using Bonferroni correction. Triangles represent estimated pandemic correlations. Red color indicates significant deviation from pre-pandemic correlation. Correlation is highest for all-cause and is more pronounced during the pandemic period (red triangle), which suggests a direct impact of the virus on these conditions.

**Figure S13.**
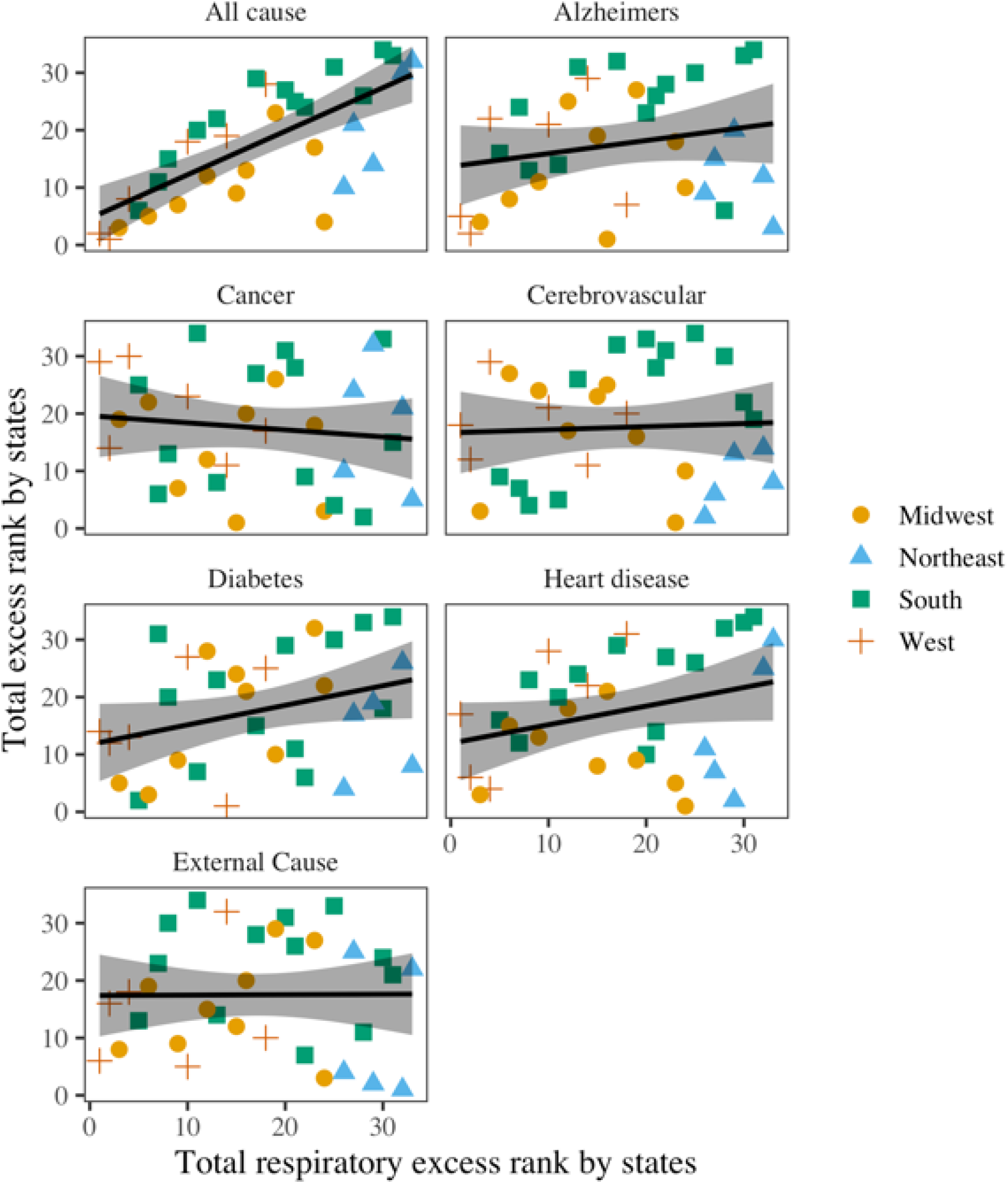
Rank correlation between total COVID-19 mortality and total excess mortality for other causes, across 33 states. Black lines represent the best fit regression lines. Shaded areas represent the 95% confidence intervals. The states have been categorized into the Midwest, Northeast, South, and West. Respiratory deaths are moderately to highly correlated with all-cause (rho = 0.73, 95% CI: 0.47 - 0.90)).

**Figure S14:**
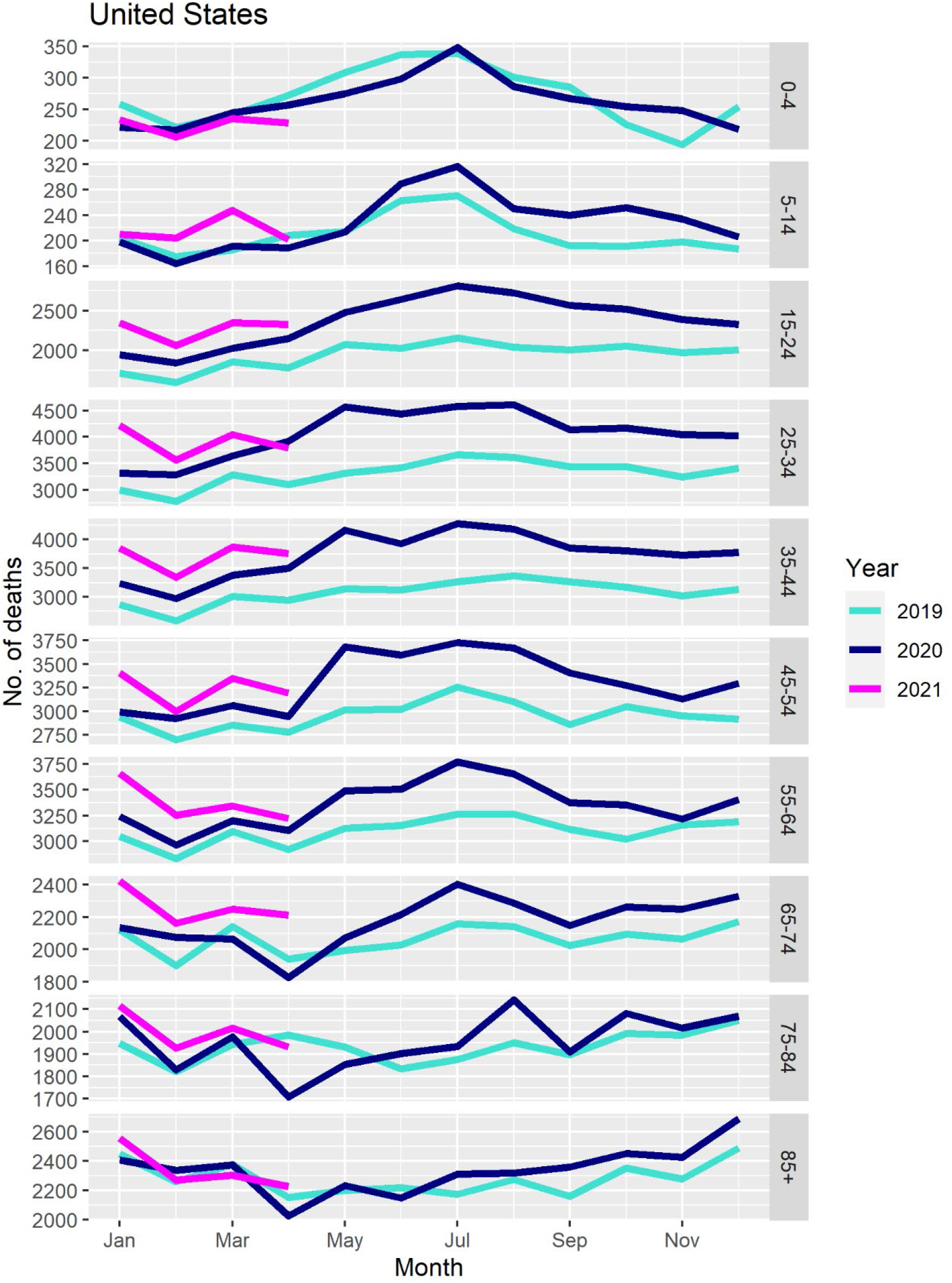
Monthly number of deaths from external causes by age group, US, January 2019-April 2021.

**Supplementary Table 1:** Evaluating the direct and indirect impact of the COVID19 pandemic nationally. Regression of national cause-specific weekly excess deaths against interventions and COVID-19 intensity (60 pandemic weeks).

| Cause of Death | Univariate |  | Multivariate |  |
| --- | --- | --- | --- | --- |
|  | COVID-19 | Interventions (GRI) | COVID-19 | Interventions (GRI) |
| All Cause | 1.158 ± 0.054*** | -0.013 ± 0.029 | 1.157 ± 0.054*** | -0.012 ± 0.010 |
| Alzheimer's | 0.020 ± 0.005*** | -0.001 ± 0.001 | 0.020 ± 0.005*** |  |
| Diabetes | 0.018 ± 0.003*** | 0.000 ± 0.001 | 0.018 ± 0.003*** |  |
| Cancer | -0.001 ± 0.005 | -0.004 ± 0.001** |  | -0.004 ± 0.001** |
| External Cause | -0.004 ± 0.007 | 0.007 ± 0.001*** | -0.006 ± 0.005 | 0.007 ± 0.001*** |
| Cerebrovascular | 0.008 ± 0.003** | -0.000 ± 0.001 | 0.008 ± 0.003** |  |
| Heart Disease | 0.052 ± 0.016*** | -0.003 ± 0.003 | 0.053 ± 0.016*** | -0.004 ± 0.003 |

\*Government response index (GRI) records the stringency of government.
Significance levels: 0-0.001 (\*\*\*), 0.001-0.01 (\*\*), 0.01-0.05 (\*)

**Supplementary Table 2:** Evaluating the direct and indirect impact of the COVID19 pandemic across states. Regression of state- and cause-specific cumulative excess death rates for the period March 2020-April 2021 against average interventions and COVID-19 intensity in the same period and state (33 states).

| Cause of Death | Univariate |  |  | Multivariate |  |  |
| --- | --- | --- | --- | --- | --- | --- |
|  | COVID-19 | Interventions (GRI) | Pandemic baseline | COVID-19 | Interventions (GRI) | Pandemic baseline |
| All Cause | 1.103 ± 0.258*** | -0.012 ± 0.035 | 0.112 ± 0.099 | 1.147 ± 0.260*** | -0.028 ± 0.028 |  |
| Alzheimer's | 0.025 ± 0.042 | -0.003 ± 0.004 | -0.010 ± 0.165 | 0.031 ± 0.043 | -0.003 ± 0.004 |  |
| Diabetes | 0.023 ± 0.035 | -0.003 ± 0.004 | 0.099 ± 0.285 | 0.029 ± 0.036 | -0.003 ± 0.004 |  |
| Cancer | -0.006 ± 0.087 | 0.002 ± 0.009 | 0.025 ± 0.149 | -0.006 ± 0.087 | 0.002 ± 0.009 | 0.025 ± 0.149 |
| External Cause | -0.007 ± 0.062 | -0.003 ± 0.007 | 0.196 ± 0.175 |  |  | 0.196 ± 0.175 |
| Cerebrovascular | 0.000 ± 0.044 | -0.002 ± 0.005 | -0.017 ± 0.210 |  | -0.002 ± 0.005 |  |
| Heart Disease | 0.092 ± 0.100 | -0.004 ± 0.011 | 0.037 ± 0.100 | 0.092 ± 0.100 |  |  |

\*Government response index (GRI) records the stringency of government interventions. (state cumulative)
Significance levels: 0-0.001 (\*\*\*), 0.001-0.01 (\*\*), 0.01-0.05 (\*)

**Supplementary Table 3:** Evaluating the direct and indirect impact of the COVID19 pandemic by age group. Regression of national age-specific weekly all-cause excess deaths against interventions and COVID-19 intensity (60 pandemic weeks).

| Age Group | Univariate |  | Multivariate |  |
| --- | --- | --- | --- | --- |
|  | COVID-19 | Interventions (GRI) | COVID-19 | Interventions (GRI) |
| Under 25 years | $0.004 \pm 0.002^*$ | $5.955 \pm 0.880^{***}$ | | $5.235 \pm 1.534^{**}$ |
| 25-44 years | $0.038 \pm 0.005^{***}$ | $28.80 \pm 2.610^{***}$ | $0.023 \pm 0.003^{***}$ | $22.49 \pm 2.142^{***}$ |
| 45-64 years | $0.199 \pm 0.010^{***}$ | $77.74 \pm 14.38^{***}$ | $0.180 \pm 0.009^{***}$ | $29.18 \pm 5.611^{***}$ |
| 65-74 years | $0.264 \pm 0.008^{***}$ | $73.11 \pm 20.23^{***}$ | $0.262 \pm 0.009^{***}$ | $2.486 \pm 5.625$ |
| 75-84 years | $0.320 \pm 0.015^{***}$ | $63.04 \pm 26.68$ | $0.338 \pm 0.015^{***}$ | $-27.97 \pm 9.612^{**}$ |
| 85 years and older | $0.381 \pm 0.025^{***}$ | $75.37 \pm 33.86^*$ | $0.403 \pm 0.027^{***}$ | $-33.07 \pm 17.27$ |

Significance levels: 0-0.001 (\*\*\*), 0.001-0.01 (\*\*), 0.01-0.05 (\*)

## Notes

### Competing Interest Statement

The authors have declared no competing interest.

